# BADLOCK: A Rapid, Portable, Inexpensive Diagnostic for Bacterial Pathogen and Resistance Detection in Resource-Limited Settings

**DOI:** 10.1101/2025.08.11.25332217

**Authors:** David J. Roach, Bhrus Pun Sangruji, Shriya Bhat, Selama Tesfamariam, Ishay Ben-Zion, Miriam Bern, Josephine Bagnall, Noam Shoresh, Lisa Milien, Roby P. Bhattacharyya

**Affiliations:** Division of Infectious Diseases, Brigham and Women’s Hospital, Boston, MA; Infectious Disease and Microbiome Program, Broad Institute of MIT and Harvard, Boston, MA; Tufts University, Boston, MA; Harvard University, Boston, MA; Kaiser Permanente Bernard J. Tyson School of Medicine, Pasadena, CA; Division of Infectious Diseases, Department of Medicine, Massachusetts General Hospital, Boston, MA

## Abstract

Antimicrobial resistance is a major global health threat, with disproportionate impact in regions with limited diagnostic infrastructure. To address this challenge, we developed BADLOCK (Bacterial and AMR Detection by SHERLOCK), a rapid, low-cost molecular diagnostic platform for direct detection of bacterial pathogens and resistance genes from clinical samples. BADLOCK operates as a one-pot CRISPR-Cas13a reaction capable of detecting nine bacterial species and four major resistance genes directly from positive blood culture. It requires only a heat block and supports both fluorescence and paper-based lateral flow readouts. We validated BADLOCK on a prospectively collected clinical cohort of 194 blood culture specimens, supplemented with 69 mock samples generated from banked isolates enriched for targeted resistance genes. Across all cohorts, we conducted 2,224 individual reactions, achieving 97.6% accuracy (2,171/2,224) at the reaction level. At the assay level, 89.5% (274/306) showed perfect or partial concordance with gold-standard species and resistance gene detection, including 255 assays with perfect concordance and 19 with partial concordance (correct detection of at least one pathogen). This included an evaluation of BADLOCK as a potential culture-free diagnostic for urinary tract infections (UTIs), achieving 98.0% reaction-level accuracy. At the assay level, 90.7% (41/43) were perfectly concordant with gold-standard detection of both species and resistance genes, with 2 additional assays showing partial concordance. To our knowledge, this represents the first demonstration of the CRISPR-Cas13a diagnostic platform on clinical bloodstream infections to date and supports BADLOCK’s potential as a practical and scalable solution for rapid pathogen and resistance gene detection in resource-constrained settings.

## INTRODUCTION

Antimicrobial-resistant (AMR) organisms are considered by the World Health Organization (WHO) to be one of the ten greatest healthcare threats facing humanity^1^. The burden of bacterial infections and AMR is most severe in low- and middle-income countries (LMICs), where the medical infrastructure necessary for their effective diagnosis and treatment is lacking^2–5^. In 2019 alone, there were an estimated 1.27 million global deaths due to resistant bacterial infections^2^, and this is projected to reach over 8 million associated deaths annually by 2050^6^, resulting in increasing calls to improve diagnostic strategies for containing the epidemic^7–9^. One of the most concerning clinical manifestations of AMR is in bloodstream infections (BSIs), which are among the most severe and life-threatening infectious syndromes^10,11^. BSIs can be caused by a wide range of bacterial pathogens, but the mortality rate is significantly higher when resistant bacteria are involved^10,12,13^, and the early identification of these pathogens can improve patient survival^14–16^. Therefore, a low-cost method to rapidly detect bacterial species and their AMR genes in clinical samples would be an invaluable tool for clinicians.

The standard diagnostic workflow for BSIs involves several sequential steps, each requiring substantial time, specialized infrastructure, and trained personnel (Figure 1a). In brief, blood from a patient with suspected bacteremia is inoculated into liquid culture media and incubated until microbial growth is detected. Positive blood cultures are then sub-cultured onto solid media, where individual bacterial colonies are isolated for species identification. In well-resourced laboratories, this identification is typically performed using advanced mass spectrometry-based techniques, such as MALDI-TOF (Matrix-Assisted Laser Desorption/Ionization - Time of Flight), while in lower-resource settings, traditional biochemical testing remains the primary method. Following species identification, antimicrobial susceptibility testing (AST) is conducted either with automated systems that assess growth inhibition in the presence of antibiotics or with manual methods such as broth microdilution, Kirby- Bauer disc diffusion or E-testing^17^. Depending on bacterial growth rate, the availability of resources, and laboratory workflow efficiency, this entire process can take anywhere from 2 to 7 days^18^, potentially delaying the initiation of targeted antimicrobial therapy.

**Figure 1.**
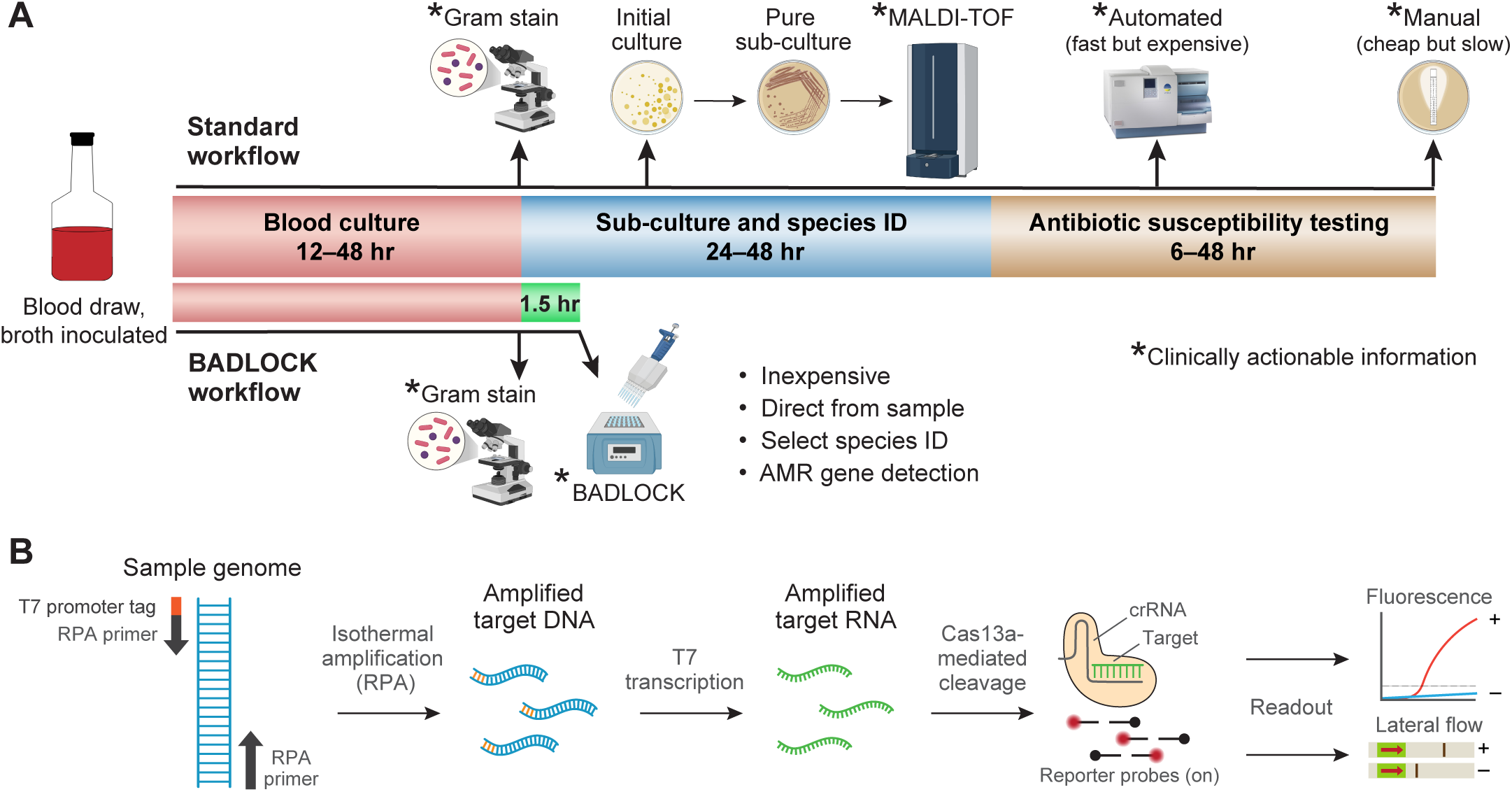
(A) Schematic representation of a standard clinical microbiology workflow for positive blood cultures in a well-resourced laboratory (top), demonstrating multiple sequential steps, extensive hands-on time, and reliance on complex instrumentation. In contrast, the BADLOCK workflow (bottom) demonstrates significantly reduced resource and instrument utilization and provides a much faster turnaround time for clinically actionable information. (B) Molecular mechanism of the SHERLOCK platform. Pre-specified gene targets are amplified isothermally using recombinase polymerase amplification (RPA). The forward primer includes a universal T7 tag, enabling transcription of the amplified gene target into RNA. The CRISPR/Cas13a complex then binds to its cognate RNA target, activating its enzymatic function and triggering trans-enzymatic cleavage. This results in the indiscriminate cleavage of nearby reporter molecules, producing a detectable signal that can be visualized as fluorescence or as a colorimetric readout on a lateral flow strip.

In recent years, a growing number of alternative approaches have emerged to address the delays inherent in traditional workflows, including rapid, gene-based diagnostic platforms capable of simultaneously detecting bacterial species and AMR genes directly from cultures^19,20^. These methods have demonstrated significant improvements in time-to-appropriate antibiotic therapy and patient survival^16^. However, they can come with substantial challenges, including high costs, a large infrastructural footprint, and the need for sophisticated laboratory facilities and specialized technical training. As a result, these advanced diagnostics are often inaccessible in under-resourced regions, which disproportionately bear the global burden of bacterial infections and associated mortality^2^. This underscores the urgent need to develop rapid, low-cost bacterial diagnostic tools specifically designed for deployment in resource-limited settings^6,17,21^.

Emerging nucleic acid detection technologies, such as SHERLOCK (Specific High-Sensitivity Enzymatic Reporter Unlocking), offer promising solutions for addressing this challenge^22,23^. SHERLOCK is a low-cost, portable, and highly programmable CRISPR/Cas13a-based system that enables the sensitive detection of nucleic acid sequences, including those from human pathogens^24–26^. Its minimal laboratory infrastructure requirements and compatibility with lateral flow testing (LFT) assays make it particularly well-suited for deployment in low-resource environments^24^. Leveraging the CRISPR/Cas13a enzymatic platform (Figure 1b), we have developed a novel diagnostic assay, BADLOCK (<u>B</u>acterial and <u>A</u>MR <u>D</u>etection by SHER<u>LOCK</u>), which enables the rapid identification of diverse bacterial species and key AMR genes directly from blood cultures. This approach streamlines the standard clinical microbiology workflow, reducing both the time and complexity required for pathogen identification while simultaneously providing critical information on clinically relevant gene content to guide targeted therapy. Designed in alignment with World Health Organization (WHO) recommendations for diagnostics in low-resource settings^21^, BADLOCK is portable, fast, low-cost, and requires minimal infrastructure, offering a practical and impactful solution for use in LMICs. In this proof-of-concept study, we demonstrate the capability of BADLOCK to accurately identify nine different gram-negative rod (GNR) bacterial species directly from clinical blood cultures, which together account for over 3.3 million global deaths annually^3^ and represent more than 80% of GNR bloodstream infections within our institution. We additionally demonstrate its ability to detect major epidemic resistance genes, underscoring its potential to address critical diagnostic gaps in global health. Finally, we demonstrate that its flexible and robust format makes it a promising diagnostic tool for other biospecimens beyond blood cultures.

## RESULTS

### Assay target development

Bloodstream infections (BSIs) can be caused by a wide variety of highly diverse bacterial species^11^, posing significant challenges for the development of comprehensive gene-based diagnostics. To enhance our assay’s practicality and clinical utility, we focused on species that collectively account for ∼80% of gram-negative BSIs within our local institution (Figure S1), which included *Escherichia coli*, *Klebsiella pneumoniae*, *Enterobacter cloacae* complex, *Citrobacter freundii*, *Serratia marcescens*, *Proteus mirabilis*, *Pseudomonas aeruginosa,* and *Acinetobacter baumannii*. We also included *Stenotrophomonas maltophilia* due to its intrinsic resistance to many commonly used antibiotics and unique treatment requirements that would affect clinical management^27^. For the resistance gene panel, we targeted widespread beta-lactamase genes that are strongly associated with resistance to broad-spectrum beta-lactams. These genes are linked to strains that are difficult to treat^27^, resistant to many empiric regimens, and exhibit higher mortality^13,28^. Specifically, we targeted the extended-spectrum beta-lactamase gene *bla*_­_and the carbapenemase genes *bla*_­_, *bla*_­_and *bla*_­_. We elected to forego *bla*_­_and *bla*_­_due to their lower prevalence both globally^29^ and within our study population^30,31^. We employed three distinct strategies for target development: (i) targeting individual pathogenic species; (ii) detecting clusters of closely related species with similar clinical significance, such as the *Enterobacter cloacae* complex, thereby minimizing the overall panel size; and (iii) identifying conserved regions within clinically relevant antimicrobial resistance (AMR) gene families^26^ (Figure 2).

**Figure 2.**
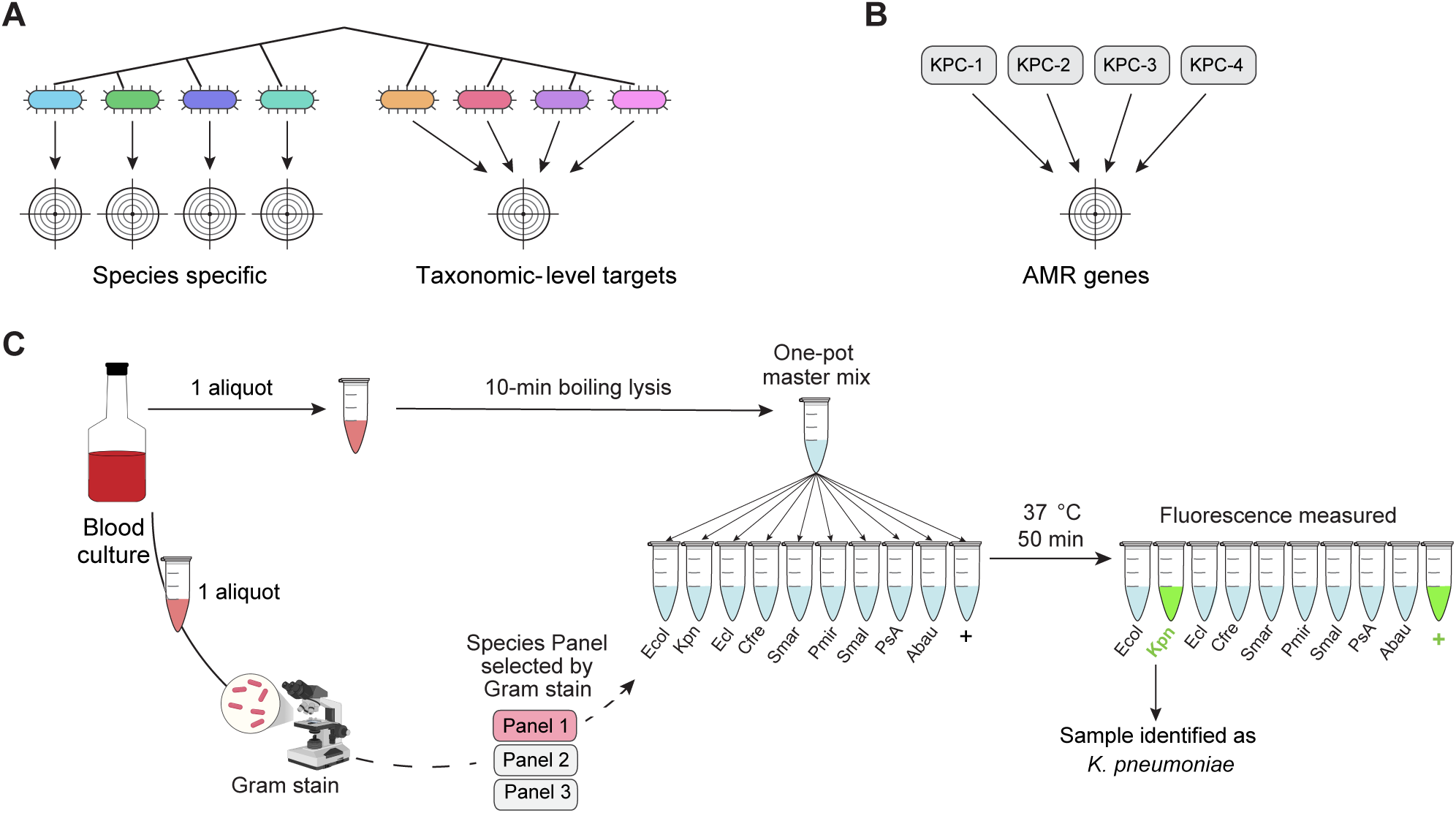
(A) Overview of the approach to species primer and guide development, showing that certain targets map to individual species, whereas others capture a group of closely related bacteria with similar clinical management pathways. (B) AMR gene targets are designed to capture most sequence variants for a given gene by targeting conserved regions. (C) BADLOCK workflow. Once blood cultures turn positive, a heat lysis step is performed while a Gram stain is concurrently conducted. Gene panel selection is guided by gram-stain morphology, with this study focusing exclusively on gram-negative rods. A single master mix is prepared and divided among different RPA primer and CRISPR guide pairs, each specifying a unique target. The reactions are incubated for 50 minutes, after which fluorescence values are measured to determine target detection. For simplicity, only gram-negative species targets are shown here.

The target design process has two layers of specificity, which include an isothermal amplification of a target region using recombinase polymerase amplification (RPA) combined with a 28bp CRISPR-Cas13a target within the amplicon (Figure 1b). Our initial approach focused on the topoisomerase I gene for species-specific targets, as it was identified through a genome-wide scan to be highly conserved within species yet sufficiently divergent between them to enable precise detection with CRISPR guides^26^. Using this strategy, we developed targets for 4 species but were unable to successfully amplify them with RPA in 3 others. In those, we identified genes from the literature that are conserved within and unique to the species of interest^32–34^ (Table S1). In *E. coli*, we encountered a unique challenge, since the RPA reagents are produced in the laboratory strain *E. coli* K12, leading to false positives. As a solution, we targeted the *chuA* gene, which is present in 80-90% of pathogenic *E. coli* strains but is absent from *E. coli* K12, a strategy used by other molecular diagnostic approaches using RPA to detect *E. coli*^35,36^. To detect the six species that constitute the taxonomic grouping called the *Enterobacter cloacae* complex^37^, we adapted a novel computational algorithm to identify conserved sequences shared within these species but absent from non-target species (Methods). For AMR gene detection, we utilized previously identified conserved regions within the genes of interest^26^. After identifying putative targets, we utilized ADAPT^38^, a computational tool designed for CRISPR guide selection, and subsequently designed flanking RPA primers designed to amplify DNA segments containing the CRISPR target sites.

Each primer and guide combination was first tested on genomic DNA from at least two banked isolates per species of interest to verify functionality and specificity. This process was iterative, with primer and guide combinations refined as needed to ensure specificity and reproducibility. Using this approach, we successfully developed RPA primers and CRISPR guide targets for all species of interest (Table S1). To identify targets within the select AMR genes, we designed RPA primers around known CRISPR guides^26^ but found that the specificity for *bla*_­_was low using the previously identified guide, requiring a guide redesign using ADAPT.

### One-pot assay design and optimization for use on blood cultures

To optimize the assay for clinical microbiology laboratory workflows, where minimizing hands-on technician time is critical, we developed a flexible one-pot approach that integrates RPA^39^ with Cas13-based detection in a single reaction. While similar approaches have utilized freeze-dried RPA pellets followed by master mix preparation^25^, we found this method to be less flexible to varying sample input numbers. Instead, we adopted a liquid-based RPA format as our starting point, but initial experiments revealed that the standard liquid-based RPA reaction yielded less robust amplification when combined with the Cas13 detection buffers. To address this, we systematically varied buffer formulations, testing different relative concentrations and salt contents to identify an optimal mix. The best-performing formulation that maintained the final reaction volume used magnesium acetate from the RPA reaction as the sole magnesium source, incorporated desalted reaction buffers, and utilized an RPA buffer at 0.66x the manufacturer’s recommended concentration (see Methods).

We next sought to transition from genomic DNA to bacterial cultures in laboratory media as the starting template for our assay. While detergent-based lysis methods have previously been used^40^, we opted for a simple heat lysis step to simplify the workflow. The lower limit of detection (LOD) for broth cultures was determined to be 10^3^ colony-forming units (CFU) per microliter for three representative species (Figure S2). Importantly, the intended target biospecimen for our assay is positive blood cultures, which typically contain bacterial concentrations between 10^6^ and 10^8^ CFU/mL. Although bacterial cultures from LB media could be directly loaded into a reaction with reliable results, initial attempts to use undiluted blood culture specimens yielded low signal, likely due to interference from blood components within the sample matrix^39^. To address this issue, we performed serial dilutions and identified that a 1:10 dilution of the blood culture reliably produced positive results.

To establish robust thresholds for positivity for each species target in clinical specimens, we used a previously validated approach^41^, defining the threshold as six times the standard deviation above the mean RFU measured for each CRISPR target in the absence of its corresponding template. Thresholds were calculated using 24 off-target clinical samples for the species targets and 12 off-target banked samples for the AMR gene targets (Figure S3). To ensure assay viability, a positive control was created using a single-stranded synthetic DNA sequence not predicted by BLAST^42^ to be in any bacteria (Figure 2c and Supplementary Table S1). Each BADLOCK assay is composed of a panel of different gene targets that are run in parallel reactions (Figure 2c; Figure 3).

**Figure 3.**
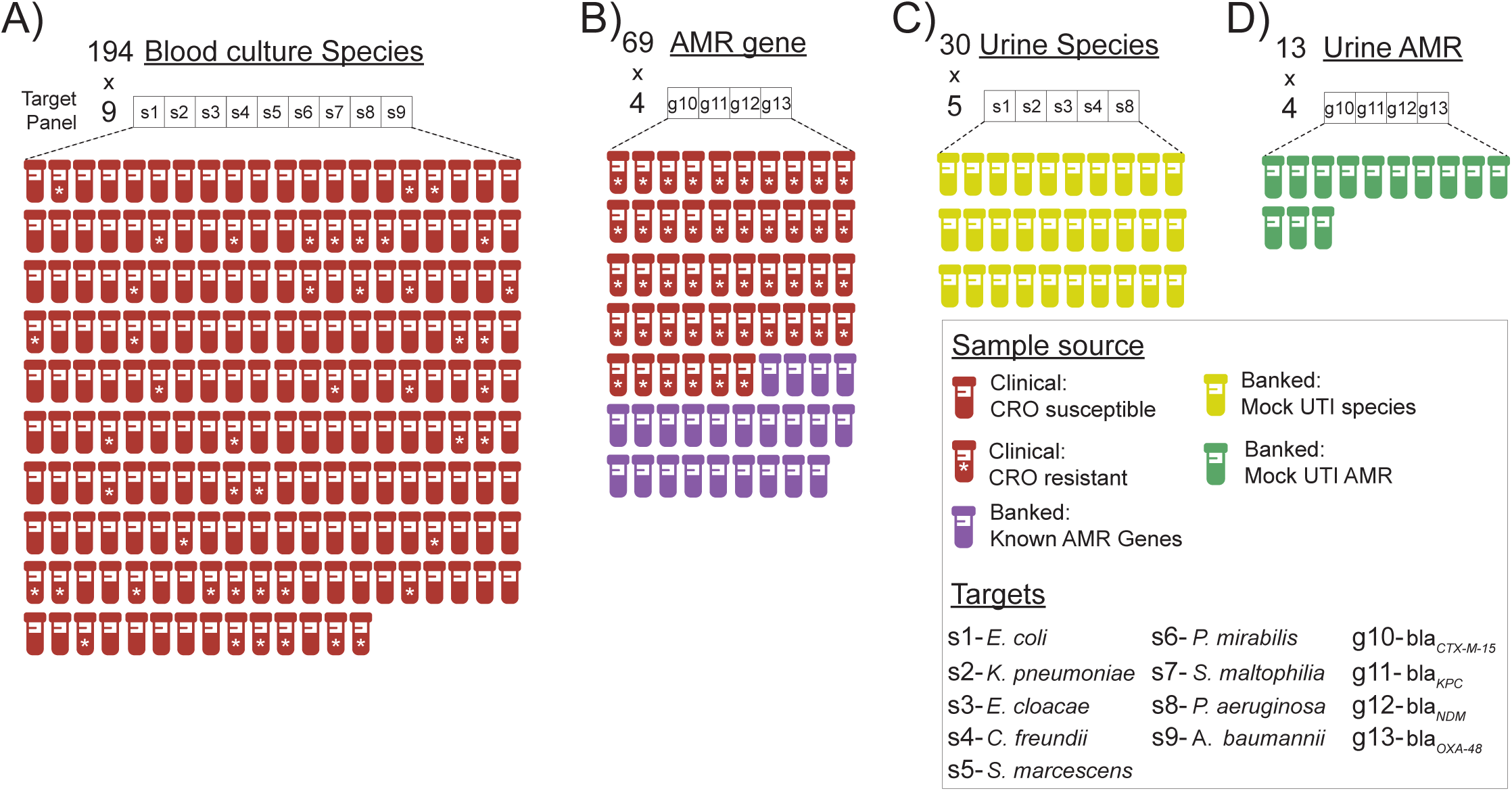
Overview of BADLOCK testing cohorts and associated CRISPR targets. Summary of each experimental group evaluated in this study, including A) species testing on clinical blood culture specimens; B) AMR gene testing on both clinical samples and banked laboratory isolates; C) species testing on mock UTI samples created from banked isolates; and D) AMR testing on mock UTI samples also created from banked isolates. Each cohort is annotated with the specific CRISPR targets used, illustrating the number and distribution of species- and resistance gene-level detection across study phases. Each sample is tested for all species targets within the target panel simultaneously, which are represented in the boxes above each cohort. “CRO” is the abbreviation of the 3^rd^ generation cephalosporin ceftriaxone.

To maintain consistency in terminology, we define an “assay” as the complete panel of targets run simultaneously, and a “reaction” as an individual target within that panel. We use “false positive” and “false negative” to describe miscalls at the reaction level. At the assay level, we define “perfect concordance” as cases where all reactions within the assay were called in agreement with the clinical laboratory results, “partial concordance” as cases where at least one true positive was detected alongside one or more miscalls, and “discordant” as cases in which no true positives were detected or, in the case of off-panel species, when at least one false positive was called. In alignment with FDA guidance on reporting diagnostic results for novel assays^43^ and consistent with recently published evaluations for other multiplex panels^44^, we report performance using positive percent agreement (PPA) and negative percent agreement (NPA). These metrics, analogous to sensitivity and specificity, are defined as follows: PPA is the proportion of true positives identified by the assay (concordant positives divided by all positive culture results), and NPA is the proportion of true negatives identified (concordant negatives divided by all negative culture results).

### BADLOCK’s species identification performance on a clinical cohort

We next evaluated BADLOCK on a cohort of frozen aliquots from 194 consecutive positive blood cultures identified as gram-negative rods, collected by the Massachusetts General Hospital (MGH) clinical microbiology laboratory (Figure 3). These samples had undergone the standard clinical microbiology workflow, including Gram-stain morphology, bacterial species identification, and antimicrobial resistance testing. Among the cohort, 157 specimens matched at least one species included in our assay panel, while 37 specimens contained exclusively off-target species (Figure S1). Additionally, we specifically wanted to interrogate polymicrobial samples (that is, when more than one species are causing bacteremia and are grown from culture) as this is a known challenge in clinical microbiology diagnostics^45^. In total, 18 specimens were polymicrobial; 8 contained one on-panel species, 6 contained two, and four contained no on-panel species.

Because each target is processed independently in separate reactions within the assay (Figure 2c), these 194 samples amounted to 1746 individual reactions; reaction-level calls were correctly made in 97.7% (1705/1746) instances across the cohort (Figure 4 and Figure S4). Among the 158 samples containing species targeted by BADLOCK, at least one target was correctly identified in 88.0% (139/158) of cases. Across the cohort on initial testing, there was perfect or partial concordance with subculture and MALDI-TOF in 88.1% (171/194) of cases, with 157 assays (80.9%) demonstrating perfect concordance and 14 (7.2%) with partial concordance. There were a total of 19 false positive reactions (across 17 samples) and 22 false negatives, with two samples containing both (Figure S4). BADLOCK identified at least one species in 100% (6/6) of polymicrobial cultures containing multiple on-panel species and both species in 50% (3/6) of cases. It identified 62.5% (5/8) in those polymicrobial samples containing a single on-panel species. In the 37 samples containing only off-panel species, BADLOCK detected no organisms in 34 (91.9%), indicating selectivity for intended targets (Figure S4). Individual performance characteristics for each RPA primer/CRISPR guide combination within the clinical cohort were calculated and varied across targets, with PPA ranging from 60% (*S. maltophilia*) to 100% (*P. aeruginosa* and *P. mirabilis*). The NPA, conversely, was high across the cohort, ranging from 97.7% to 100% across species targets (Figure 4 and Table S2).

**Figure 4.**
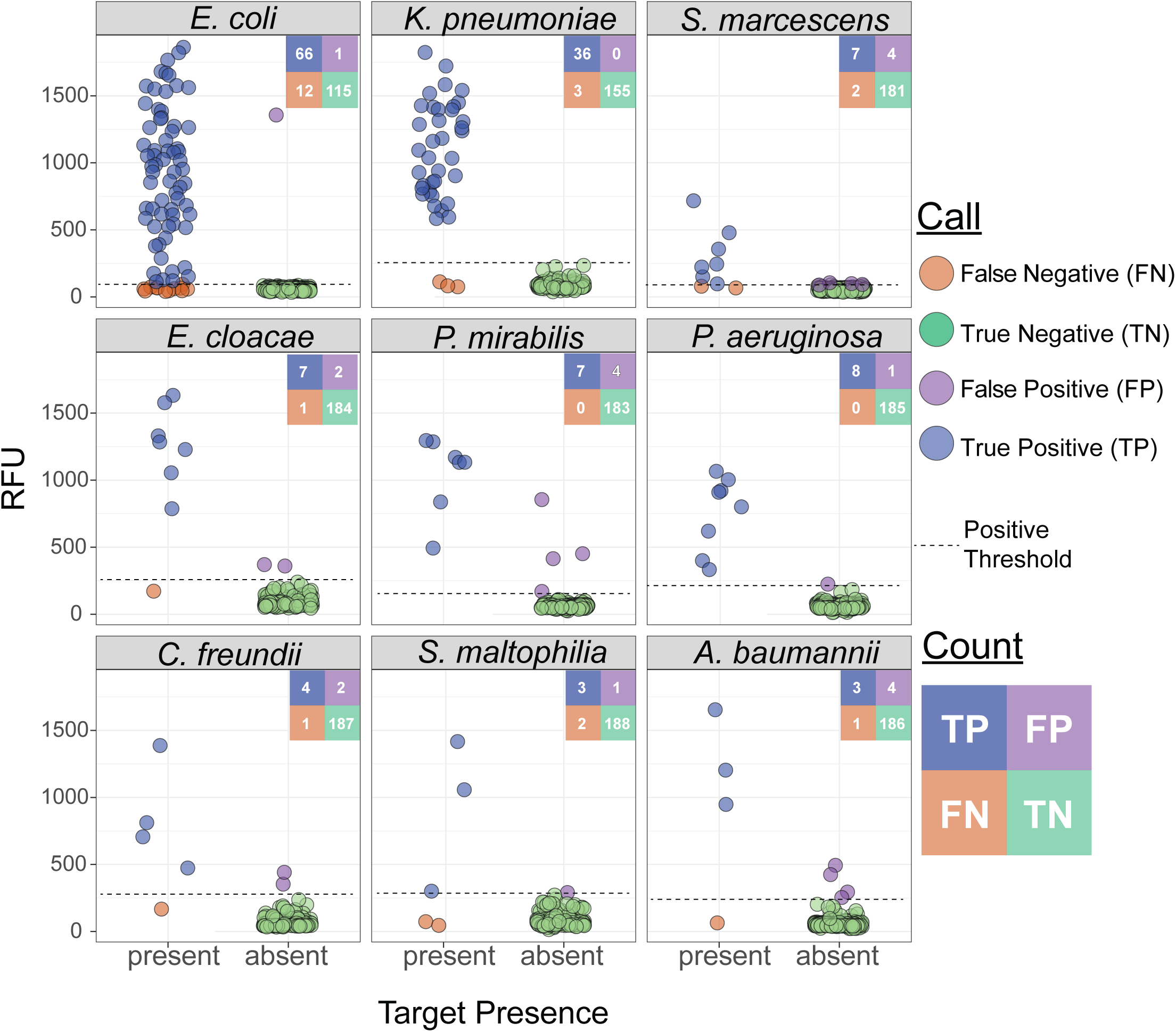
Performance of BADLOCK species panel on clinical positive blood culture samples. Each panel displays a scatterplot of RFU values for the corresponding species target across all 194 samples. A confusion matrix summarizing the total counts for each call is included for each species. Ground truth (x-axis) was determined by routine testing in the clinical microbiology laboratory using the MALDI-TOF platform.

### Performance on antimicrobial resistance gene testing

To optimize reagent usage in this pilot study, we focused our testing on clinical samples known to be resistant to third-generation cephalosporins (which are more likely to harbor the ESBL or carbapenemase genes targeted by our assay), resulting in a final testing cohort of 46 clinical samples (Figure 3b), only one of which was carbapenem-resistant. PCR was performed for all resistance gene targets in the ceftriaxone-resistant cohort to establish a ground truth for gene presence, and long-read whole-genome sequencing was conducted on a subset of nine samples for further validation. We achieved 96.2% accuracy (177/184) at the reaction level, and perfect concordance in 84.8% (39/46) of assays, with no partially concordant assays and 15.2% (7/46) discordance. We had 100% PPA for all AMR gene targets and 100% NPA for all carbapenemase genes but notably had 7 false-positive reactions for the *bla_­_*gene target (Figure 5 and Figure S5, left panel).

**Figure 5.**
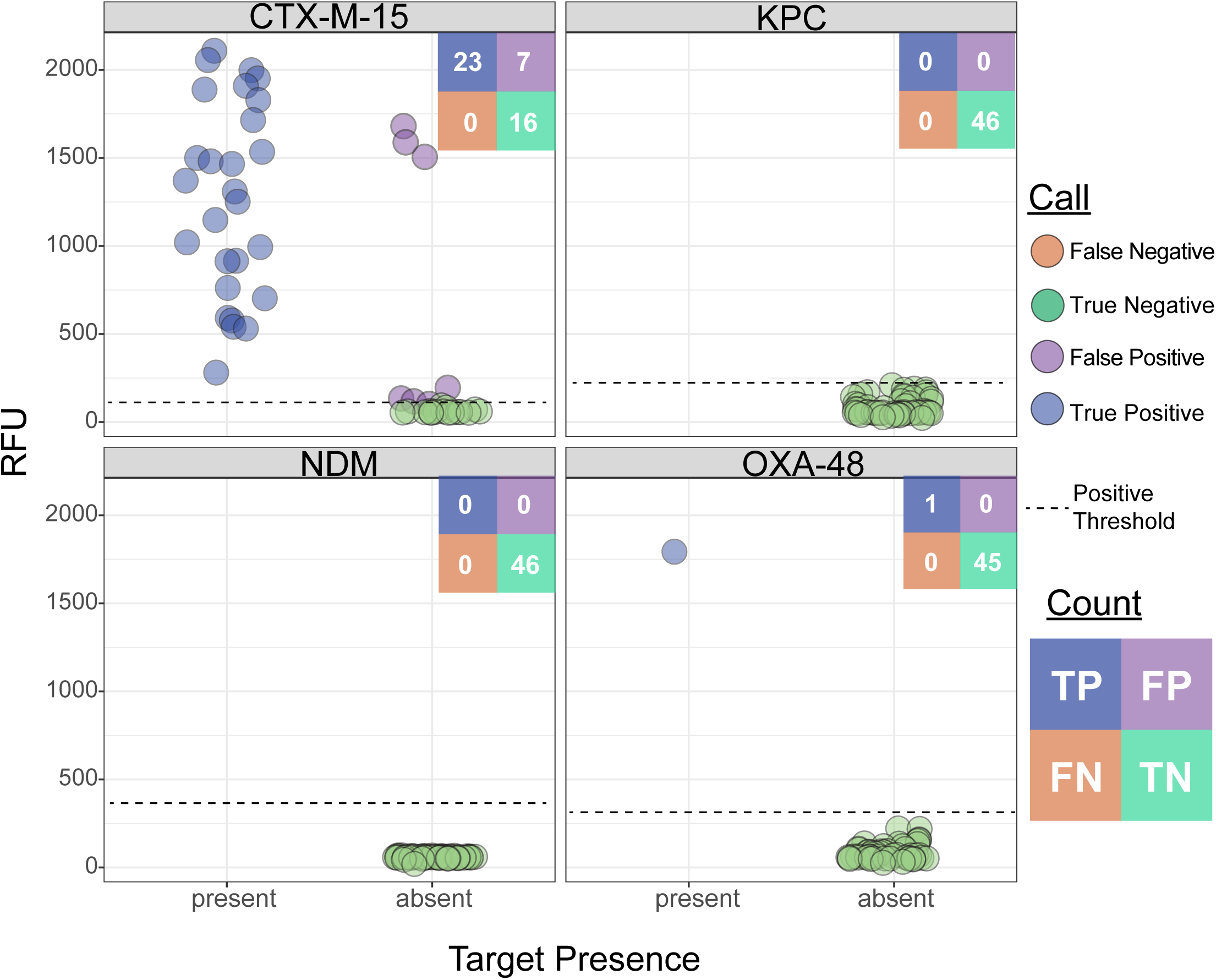
BADLOCK AMR gene testing in clinical cohort of positive blood culture specimens, limited to those 46 samples resistant to ceftriaxone, with gene content (x-axis) validated by PCR testing or whole-genome sequencing.

Given the low prevalence of carbapenem resistance in the MGH clinical population^15^, we supplemented our cohort with 23 banked carbapenem-resistant isolates with known beta-lactamase content (Figure 3b and Table S3), identified through prior sequencing efforts^46^. Within this cohort of banked strains, BADLOCK had a correct reaction-level call in 98.9% (90/92) cases, resulting in perfect concordance in 91.3% (21/23) of assays and partial concordance in the remainder. The only two miscalls were from two false positives just above the threshold line (Figure 6). Importantly, nine samples contained two on-panel genes, and BADLOCK correctly identified both genes in 100% (9/9) of cases (Figure S5, right panel).

**Figure 6.**
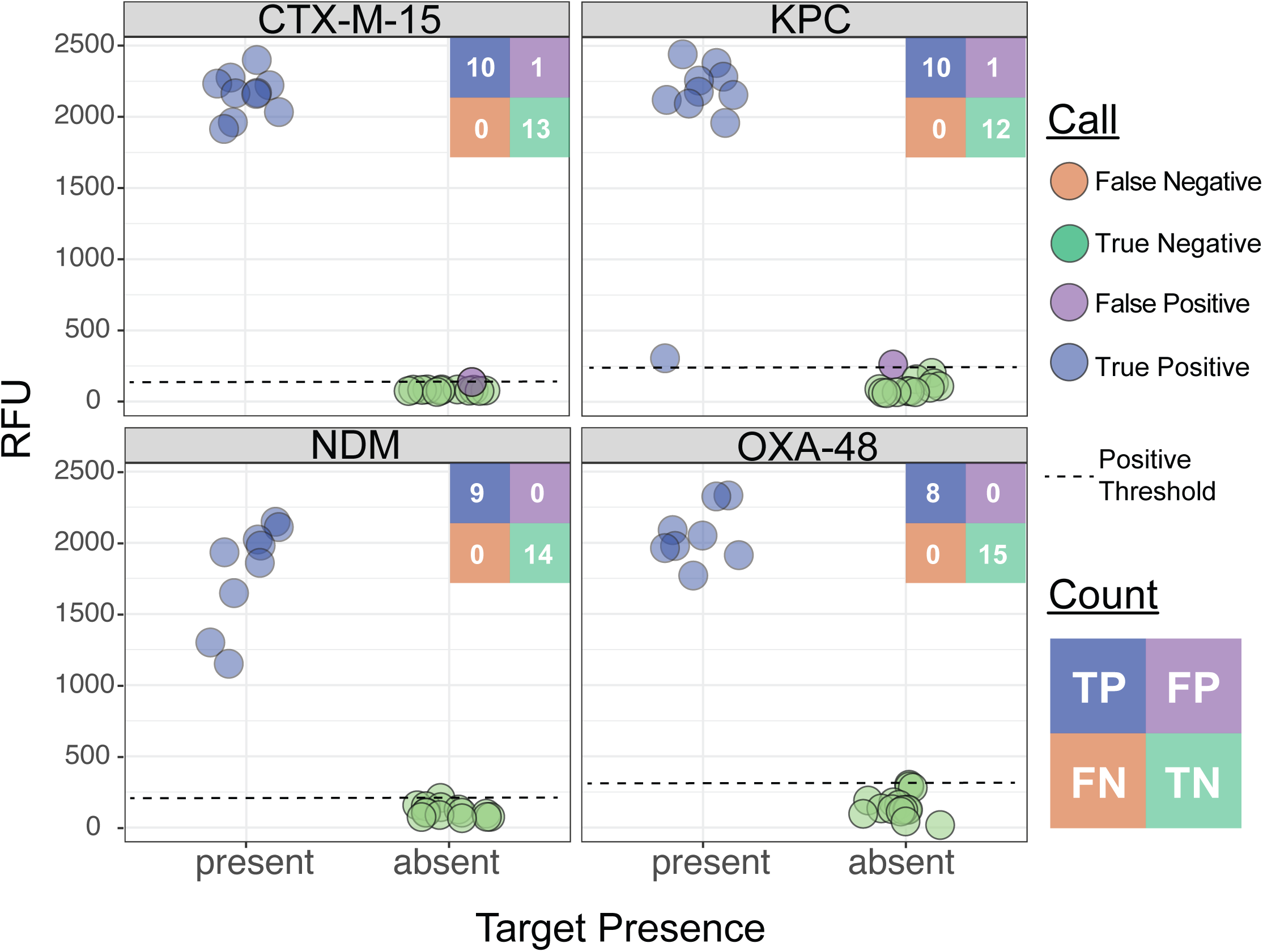
BADLOCK AMR gene panel testing performance from banked clinical laboratory strains with known genetic backgrounds, validated by whole-genome sequencing.

### Investigation of Discrepancies

We identified three distinct error modes contributing to discrepancies: (i) false negatives, where BADLOCK was negative but the gold-standard test was positive; (ii) false negatives due to absence of the molecular target, as seen with the *chuA* gene in *E. coli*; and (iii) false positives, where BADLOCK was positive but the gold-standard test was negative. To investigate these, we first repeated the assay for all discordant results in the species testing. In 43.9% (18/41) of cases, this repetition resolved the discrepancy; in the remainder, the discordant call persisted (Figures S6 and S7).

*False negatives:* Of the 22 total discordant negatives, 12 were in *E. coli*, where we had to target a non- conserved gene (*chuA*) that is not present in the laboratory strain *E. coli* K12 used to make the RPA reagents. As such, we anticipated ∼15% of our *E. coli* samples to be falsely negative^36^, which was consistent with our final detection rate of 84.6% (66/78). Of the 12 discordant negative *E. coli* samples, PCR confirmed that the *chuA* gene was absent in 11. In the remaining case (sample 106), where *chuA* was present, repeat testing yielded a robust positive result (Figure S6). The remaining 45.5 percent (10/22) of discordant negatives were in non-*E. coli* species, and three of these resolved with a single repeat run (Figure S6). In two cases, the samples were polymicrobial, potentially indicating low-abundance species within the sample (where the blood culture may have signaled positive due to another species).

*False positives:* Discordant positives were most frequently observed in *A. baumannii*, *S. marcescens*, and *P. mirabilis*, with each species accounting for four cases. *A. baumannii* had previously been used as a positive control, raising the possibility of amplicon contamination from prior runs as the source of these false-positive calls. This suspicion was supported by the fact that all *A. baumannii* discordant positives resolved when the assay was repeated at a later time point with fresh reagents (Figure S7). In all four *S. marcescens* false positives, and in the *P. aeruginosa* and *S. maltophilia* miscalls, the RFU values were just above the threshold.

All discrepancies but one resolved upon repeat testing, with the single persistent false positive for *S. marcescens* (sample 195) remaining positive but right at the threshold (see *Threshold Optimization* section below).

All seven discordant AMR results in the clinical sample cohort were false positives for *bla_­_*. Of these, three had very high RFU values, while four were only marginally above the positivity threshold (Figure 5). Two of the three high-RFU samples fully corrected on repeat testing, while one showed reduced signal but remained just above the threshold (Figure S8). Notably, all three of these samples had been batched and run together during the initial testing, raising suspicion of a cross-contamination event. For the AMR gene testing on banked isolates, the two discordant calls were a single false positive for *bla_­_*and one for *bla_­_*, both of which occurred right at the threshold and corrected upon repeat testing (Figure S8).

### Threshold Optimization

We set the initial positivity thresholds at 6 standard deviations above the mean of RFU values from off-target samples run prior to this study, but as more data was generated after running the entire sample set, we ran a *post hoc* receiver-operator curve (ROC) analysis to optimize thresholds within our cohort (Figure S9). ROC curves help identify optimal diagnostic thresholds by visualizing the trade-off between sensitivity and specificity across all possible cutoff values, enabling flexible threshold selection tailored to each diagnostic test^47^. We evaluated the performance of each species- and gene-specific assay, comparing RFU values against known true-positive and true-negative classifications. Prior to analysis, we excluded false positive samples suspected to be caused by contamination in the initial reaction, defined as those with repeat RFU values more than three- fold lower than the initial run (Figure S7 and S8), as well as *E. coli* samples lacking the *chuA* gene, as these would confound a ROC analysis (i.e., samples lacking the molecular target should not be used to set optimal thresholds for evaluating performance). Most targets demonstrated excellent diagnostic accuracy, with area under the curve (AUC) values ≥ 0.94 for all but one target. These assays showed near-perfect sensitivity and specificity at their respective optimal thresholds, as defined by Youden’s Index^48^. In contrast, the *S. maltophilia* assay had lower overall discriminative ability (AUC = 0.72) due to limited sensitivity despite perfect specificity.

### Trial on Lateral Flow Strips

CRISPR-Cas-based diagnostics offer the advantage of lateral flow compatibility^24^, presenting a promising opportunity for low-resource and point-of-care applications. However, the HybriDetect lateral flow system, which is the most commonly used platform to demonstrate this functionality, was designed for hybridization- based detection of DNA targets as opposed to enzymatic cleavage detection and was co-opted for use with CRISPR-Cas systems^23,41^, raising questions about its performance characteristics. To evaluate this, we tested the lateral flow assay on a subset of samples. Some cases exhibited a clear distinction between positive and negative signals despite ubiquitous faint signals in negative strips, such as samples 001 and 152 (Figure S10, panels A & B) and the AMR genes (Figure S11, all). However, in other instances, such as for sample 016 and the off-panel sample 040 (Figure S10, panels C & D), the faint signal in negative strips complicated interpretation, making it challenging to reliably distinguish true from false positives. These findings highlight both the potential and the current limitations of integrating CRISPR-based detection with lateral flow readouts, underscoring the need for lateral flow strips purpose-built for SHERLOCK-based platforms and further optimization to improve specificity and robustness across diverse targets.

### Proof-of-concept test on mock urinary tract infections

Urinary tract infections (UTIs) have higher concentrations of bacteria than BSIs, potentially removing the need for a culture step and enabling direct-from-sample detection with BADLOCK. To evaluate this, we tested a subset of targets representing common urinary pathogens, including *E. coli*, *K. pneumoniae*, *E. cloacae*, *C. freundii*, and *P. aeruginosa*. Mock UTI samples were generated by culturing individual bacterial isolates and creating a dilution series for each species in filtered and pooled human urine (Methods). In our initial tests, BADLOCK successfully detected *E. coli*, *E. cloacae*, and *C. freundii* at concentrations below the established clinical threshold for infection, 10^5^ CFU/mL^49^ (Figure S12), demonstrating promising sensitivity for direct-from- sample application. However, for *K. pneumoniae* and *P. aeruginosa*, the LOD was higher than the clinical threshold required for diagnosing infection. To improve sensitivity, we tested multiple approaches, including increasing template input volume, modifying reagent mixes, incorporating detergent-based lysis, and employing centrifugal concentration. We found that using centrifugal concentration significantly enhanced sensitivity, enabling detection of all urine species targets at 10^5^ CFU/mL. Using the updated protocol, we re- established detection thresholds for urine samples as described above (Figure S13).

We next generated mock UTI samples, including at least five replicates for each gene target (including both species and AMR genes) as well as five samples containing off-panel species, each spiked at 10^5^ CFU/mL. This resulted in a total of 43 assay runs, comprising 30 species assays and 13 AMR gene assays (Figure 3c and 3d). At the reaction level, we achieved 98.0% concordance (198/202), whereas at the assay level, we achieved perfect concordance in 90.6% assays (39/43), partial concordance in 4.7% (2/43), and discordant results in 4.7% (2/43). The PPA was 100% for all targets except *K. pneumoniae* and *bla*_­_, which each had a PPA of 80% (corresponding to a single false negative each). The NPA was similarly high, with 100% for all targets except *E. cloacae* and *bla*_­_, which had 96.0 and 87.5% NPA, respectively (Figure 7). Each of the four discrepancies was resolved with a single repeated run. No off-panel species yielded signal above the detection threshold, further supporting the assay’s specificity (Figure S14). This proof-of-principle shows that BADLOCK can be applied directly to a urine matrix at clinically relevant bacterial loads to reliably identify both bacterial species and AMR genes.

**Figure 7.**
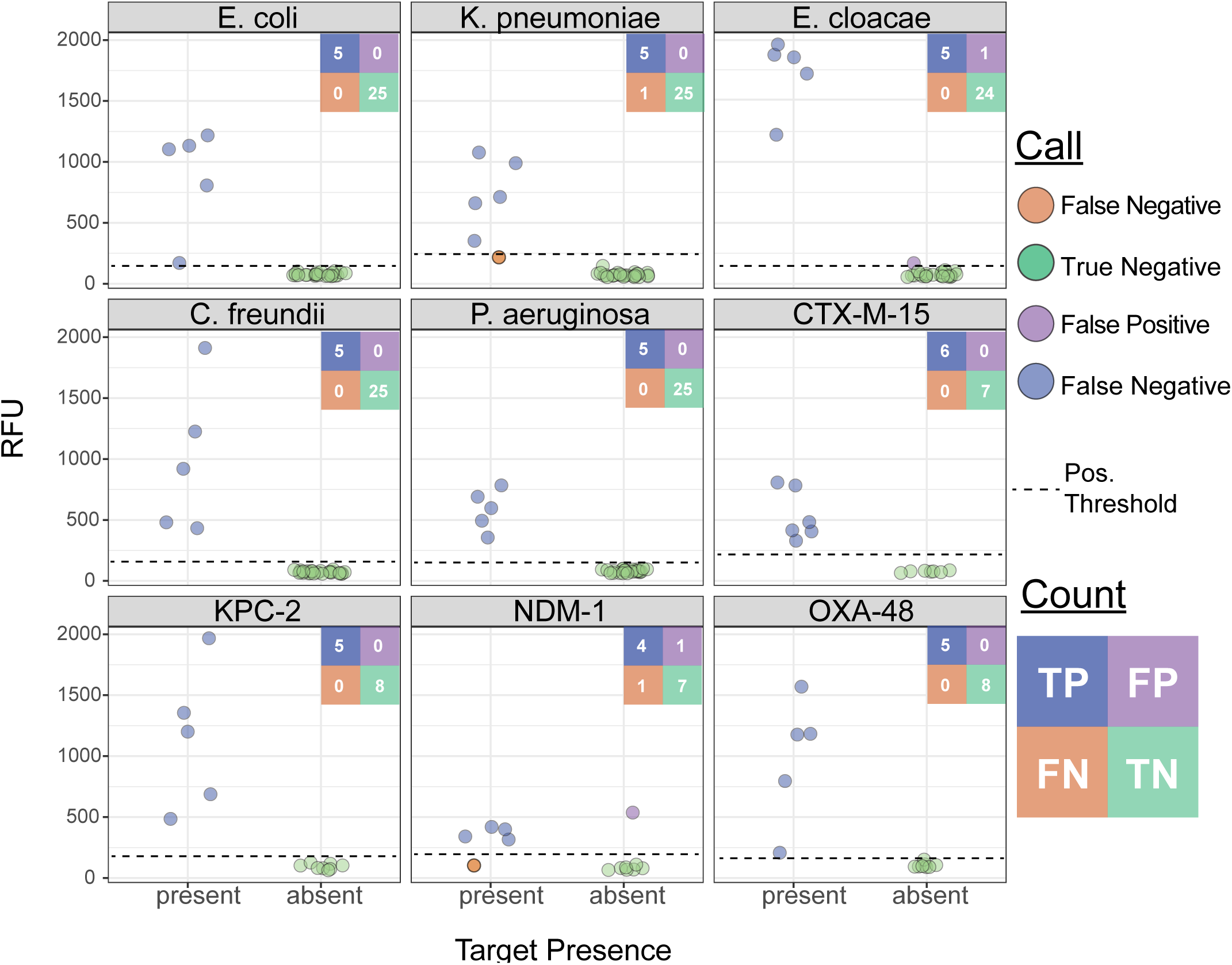
BADLOCK testing on mock urine samples, including both species and AMR gene detection.

## DISCUSSION

Here, we present BADLOCK, an inexpensive and adaptable diagnostic system designed to rapidly identify a panel of bacterial species that collectively account for over 3 million global deaths annually^3^, as well as major antimicrobial resistance determinants with significant clinical implications. We validated its utility in a retrospective study of 194 clinical blood cultures and conducted a proof-of-concept demonstration on mock UTI samples, corresponding to phase 2a and 1d studies, respectively, within a recently proposed framework for evaluating novel bacterial diagnostic platforms^17^. To our knowledge, this is the first study to evaluate a SHERLOCK-based diagnostic platform on clinical bloodstream infections, representing a critical step toward translating CRISPR diagnostics into real-world tools for the detection of clinically significant diseases. Across all cohorts and targets, we ran 2,224 individual reactions with 97.7% accuracy, reflecting the high performance of individual RPA primer and CRISPR guide combinations. Our assay-level concordance was more variable, with perfect or partial concordance ranging from 88.1% for species detection in clinical blood culture specimens to >95% in the mock UTI cohort. This distinction between overall assay concordance and reaction- level accuracy is important in modular diagnostic platforms like BADLOCK, where multiple targets are evaluated independently within a single assay and variability across targets and sample matrices can affect composite results.

While these findings demonstrate the strong potential of BADLOCK as a rapid, modular diagnostic system, several areas for improvement were identified in this initial pilot. A subset of underperforming primer and guide pairs contributed disproportionately to discordant results. Post-hoc analyses showed that thresholding can be improved with larger datasets, which helps distinguish true positives from borderline signals and reduces both false positives and false negatives. Additionally, batching during sample processing likely contributed to cross- contamination in a small number of runs, which is an issue that can be addressed with modified workflows in future versions. Finally, iterative improvements in guide selection and primer design will be key to optimizing target-level accuracy and enhancing overall assay performance.

Notably, BADLOCK employs a streamlined workflow that combines target amplification and detection in a single step, making it well-suited for integration into clinical microbiology laboratory workflows. While our testing utilized a fluorescence-based readout to support high-throughput processing, BADLOCK is also compatible with paper-based lateral flow strips; however, to fully realize its potential in low-resource settings, we advocate for the development of dedicated, clinical-grade lateral flow systems rather than repurposing of lateral flow strips designed for other applications. Such advancements could reduce infrastructure requirements to a single heat block for processing positive cultures, thereby greatly expanding access for clinical microbiology.

Our findings demonstrate the potential of BADLOCK to address key challenges in diagnostic capacity for bloodstream infections, particularly in LMICs. In many LMICs, the substantial up-front investments^50^ required for clinical microbiology laboratories—combined with the need for specialized and highly trained personnel— often limit their integration into clinical workflows. As a result, fewer than 1% of clinical laboratories have AMR testing capacity in some regions^51^. This forces reliance on syndrome-driven therapies and broad-spectrum antibiotics, contributing to inappropriate antibiotic use and accelerating antimicrobial resistance in precisely the settings that experience the most resistance^2,6^. BADLOCK was specifically designed with these limitations in mind, aligning with the REASSURED criteria established by the WHO to prioritize affordability, simplicity, and minimal infrastructure requirements^21^. By providing rapid, actionable results in a near-point-of-care format, BADLOCK offers a feasible solution for improving diagnostic capacity in resource-limited settings. Its ability to deliver results with minimal infrastructure may help bridge the critical diagnostic gap in LMICs, supporting more targeted therapy and reducing reliance on the use of empiric and increasingly ineffective antibiotics.

While the ideal diagnostic for bloodstream infections would eliminate the need for a culture step entirely, the low bacterial load typically present in these infections poses a significant challenge that has proven exceedingly difficult to overcome^52,53^. Recently developed commercial multiplex PCR assays such as the BIOFIRE® Blood Culture Identification 2 (BCID2) Panel (bioMérieux)^20^ and the VERIGENE® Blood Culture Nucleic Acid Tests (Luminex)^54^ target a similar panel of organisms and resistance genes, demonstrating the value of these targets, and of positive blood cultures as an assay input. However, the high per-run cost and significant infrastructure requirements of these assays make them prohibitively expensive for many hospitals, even in well-resourced settings. Notably, in a systematic literature search, we found no published reports demonstrating their implementation or evaluation in resource-limited settings. However, recent technological advancements such as the Mini-Lab^55^ and other innovations^56^ show promise for lowering the barrier for blood culture capacity in LMICs, presenting natural synergies with a platform like BADLOCK. Given its scalability, potential for integration into existing clinical microbiology workflows, and the low cost of components, we believe BADLOCK has the potential for broader adoption in both resource-limited and resource-rich clinical labs.

This study had several limitations. First, due to the technical constraint of using RPA reagents produced in a laboratory strain of *E. coli*, we had to target a gene that was missing from that laboratory strain, and thus not fully conserved within the species. Given that *E. coli* is the most common bacterial species implicated in gram- negative bloodstream infections, including in our cohort, this limitation led to an expected reduction in assay sensitivity. However, this could be addressed in future iterations through manufacturing processes that eliminate *E. coli* DNA contamination prior to commercialization, or produce reagents in a non-pathogenic species. Second, some species in our panel exhibited variable target performance due to a less robust RFU signal in positive samples, although our post hoc ROC analysis demonstrated improved threshold targets.

Future work will focus on guide design strategies to boost performance for these species and incorporate the optimized thresholds in *a priori* analysis. Third, this pilot was conducted within a single institution, which may limit the generalizability of the findings to other institutions or geographic regions. Although prior sequencing efforts have highlighted the institutional diversity of certain species in our panel^15^, future studies should include samples from multiple geographic regions to ensure broader applicability. However, we anticipate this limitation to have minimal impact, as our targets were selected to be conserved across all sequenced isolates of each species or gene family^26^. An additional limitation was probable cross-contamination leading to false-positives in 9 samples that retested negative, likely related to initial batched sample processing as mentioned above.

Future iterations will address this through process improvements, including the use of pre-made reaction master mixes. Finally, expanding the assay to incorporate a wider range of pathogens and resistance markers tailored to region-specific epidemiology could further enhance its utility in addressing the global burden of bloodstream infections.

In summary, we developed a CRISPR-Cas13a-based prototype assay, termed BADLOCK, as a promising step toward expanding rapid, low-cost diagnostic capacity in resource-limited settings. By validating the assay on a large cohort of clinical positive blood cultures, we demonstrated its potential for accurate pathogen detection and resistance characterization. Additionally, we explored its utility as a direct-from-sample diagnostic in urine, highlighting its adaptability and broader application in diagnosing infections from diverse sources. These findings underscore BADLOCK’s feasibility as a scalable, accessible diagnostic tool, addressing a critical gap in infectious disease management where rapid and affordable testing is most needed.

## METHODS

### Clinical Samples and Ethics Statement

Clinical blood culture aliquots were de-identified prior to use in the research laboratory. Study protocol was reviewed and approved by the Institutional Review Board (IRB) under Mass General Brigham IRB protocol 2015P002215.

### CRISPR guide design and RPA primer validation

CRISPR guides were selected by inputting the desired target genes into ADAPT, which generates a ranked list of putative guide sequences. Once a candidate guide was selected, RPA primers flanking the target region were designed. Multiple forward and reverse primers were synthesized, and all possible pairwise combinations were tested to identify viable primer sets. These were evaluated using target genomic DNA as the template, with water included as a no-template control. RPA reactions were set up in 0.2 mL PCR strip tubes using the TwistAmp® Liquid Basic Kit (TwistDx™). Each 250 µL reaction mix contained 10 µL of diluted and lysed bacterial template, 165 µL of TwistAmp® 2× Buffer (to yield a final 0.8× concentration), 22 µL of TwistAmp® 20× Core Reaction Mix (1× final concentration), 44 µL of TwistAmp® 10× E-Mix (1× final concentration), and 44 µL of a 4× dNTP mix (containing 2.5 µM of each dNTP; *Takara Biosciences*) to achieve a final dNTP concentration of 1 µM. Reactions were mixed gently by pipetting and incubated at 37 °C for 50 minutes.

Amplification products were purified using QIAquick® column purification (*Qiagen)* and visualized using Invitrogen™ E-Gel™ General Purpose Agarose Gels, 2% (*Thermo Fisher Scientific*), pre-cast with SYBR™ Safe DNA stain. A 15 µL aliquot from each RPA reaction was loaded directly into the wells of a 12-well E-Gel cassette alongside 7.5 µL of a 100 bp DNA ladder (*New England Biolabs*) to assess product size. Gels were run using the E-Gel™ Power Snap Electrophoresis System (*Thermo Fisher Scientific, Cat. No. G810*0) under default conditions for 10 minutes. Following RPA primer validation, DNA guides were synthesized (*Integrated DNA Technologies*) with universal T7 transcription tails and subsequently transcribed into RNA, as described elsewhere^26^. Each RPA primer and CRISPR guide pair was then tested in a one-pot reaction using target genomic DNA as the template and water as a no-template control (see BADLOCK Assay section below for reaction conditions). Primer-guide pairs that generated high on-target RFU values and minimal background signal in the water control were advanced for further validation. This process was repeated iteratively until viable primer-guide combinations were identified for all targets of interest.

### E. cloacae species complex guide design

To identify conserved sequences shared within different sub-species of the *E. cloacae* complex (ingroup) but absent from non-target species (outgroup), we adapted an algorithm from Thakku et al^41^. Briefly, for each ingroup or outgroup species, we downloaded up to 100 genomes which satisfied our criteria: complete assembly, latest version, and not anomalous. We then listed all the overlapping 100mers (e.g. 1-100, 2- 101, etc.) of the reference genome of *E. cloacae*, and aligned each 100mer to all ingroup and outgroup genomes using bowtie2^57^ with score-min = L,0,0 (i.e. exact match) for ingroup and score-min = L,0,-0.6 (i.e. up to ∼10 mismatches) for outgroup. 100mers that aligned, using these parameters, to all ingroup genomes and to none of the outgroup genomes were used downstream for guide design using ADAPT.

### Bacterial Collection and Standard Clinical Typing

Between October 2023-April 2024, all blood cultures taken as part of routine clinical care at Massachusetts General Hospital that signaled positive on the BACTEC FX system (*Becton Dickinson*, Sparks, MD) and were gram-negative rods by routine gram-staining were included. After staining, a 1mL aliquot of the culture was stored at -80° C. The remainder of the sample then underwent routine clinical processing with solid-media subculturing and species identification via MALDI-TOF mass spectrometry (VITEK MS, version 3.2 in vitro diagnostic Knowledge Base, *bioMérieux*, Durham, NC) and antimicrobial susceptibility testing (AST) performed on the VITEK 2 AST-GN81 Gram Negative Susceptibility Card (bioMérieux), with Clinical and Laboratory Standards Institute (CLSI) breakpoints from the M100-Ed31.

### BADLOCK assay

To limit reaction interference from blood products, 10ul of sample was added to 90ul of nuclease-free water. Heat-lysis of the samples was performed by boiling at 95° C for 10 minutes. Each BADLOCK run consisted of 10 individual reactions that combine RPA amplification, T7 transcription and Cas13a mediated detection of the target. To do so, a single master mix was made combining the following reagents: 13uL of the diluted and lysed bacterial template, 99ul TwistAmp® liquid basic 2x Buffer (TwistDx™) (0.8x final concentration), 13.2ul TwistAmp® liquid basic 20x Core Reaction Mix (1x final concentration), 26.4ul TwistAmp® liquid basic 10x E- mix (1x final concentration), 26.4ul of a dNTP mix containing each dNTP at 2.5uM (*Takara Biosciences*) for an aggregate dNTP concentration of 1uM, 26.4ul of salt-free cleavage buffer (40% by volume 1M tris-HCL at pH 7.5, 10% by volume 100mM DTT, 50% by volume nuclease-free H2O), Cas13a resuspended in 26.4ul of salt- free storage buffer (nuclease-free H2O with a final concentration of 50mM Tris-HCl at pH 7.5 and 5% glycerol) for a final Cas13a concentration of 45nM, 10.56ul of 25mM rNTPs (*New England Biolabs*®) for a final concentration of 1mM, 7.9uL of 50,000U/mL T7 polymerase (*New England Biolabs*®) for a final concentration of 1.5U/ul, 6.6uL of 40,000U/mL RNase inhibitor murine (*New England Biolabs*®) for a final concentration of 1U/ul, and 4.125uL of 8uM RNase Alert® Reporter V2 (*Life Technologies*™). This master mix was vigorously mixed and 18.7 microliters were split across each individual reaction well, which contained 0.5uL of 20uM RPA forward primer, 0.5uL of 20uM RPA reverse primer, and 1uL of 450nM CRISPR guide designed around the respective target of interest. Finally, 1ul of 280nM magnesium acetate was added to reaction as the sole cofactor and salt added to the mix, for a final concentration of 12.4nM. The assay was then run at 37° for 50 minutes. When run on spectrophotometry, each sample was checked at a wavelength of 490nm and the relative fluorescence units (RFUs) were recorded. Lateral flow assays were performed using the same reaction conditions; however, instead of Reporter V2, a 14-base poly-uracil oligonucleotide tagged with FAM at the 5′ end and biotin at the 3′ end was used. This reporter was added at the same concentration and volume as in the fluorescence-based assays.

### Creation of mock UTI samples and urine sample preparation

Stocks were streaked onto LB agar plates and incubated at 37 °C for 24 hours. A single colony from each plate was inoculated into LB broth and cultured at 37 °C with shaking until the optical density (O.D.) at 600 nm reached 0.250 - 0.500, as measured using a plate reader. Following growth, each bacterial culture was diluted in pooled, filtered human urine (*Innovative Research*) to a target concentration of 10⁸ CFU/mL, based on standard O.D. to CFU conversion values. This 10⁸ CFU/mL culture, prepared in a final volume of 100 µL, served as the starting point for a serial dilution series. All serial dilutions were carried out using the same pooled human urine as the diluent to maintain matrix consistency across all experimental conditions.

Expected CFU values were confirmed through plating. For samples between 10^4^ to 10^6^ CFU/mL, 1,000 µL was transferred into a 1.5mL tube and centrifuged at 21,300 × g for 1.5 minutes to pellet the cells. Following centrifugation, 980 µL of the supernatant was carefully removed to achieve a 50× concentration of the bacterial sample. The pellet was then resuspended and heat-lysed at 95 °C for 10 minutes, followed by cooling at 4 °C for 2 minutes. This template was then carried forward in the same manner as described above.

## Data Availability

All data produced in the present study are available upon reasonable request to the authors

## Acknowledgements

We would like to thank the Massachusetts General Hospital Clinical Microbiology Laboratory for all their efforts in sample collection that enabled the prospective portion of this work, as well as Jackson Lirette for his help with training DJR on the SHERLOCK assay.

## Author Contributions

DJR was responsible for experimental design, data analysis, and writing. BPS and SB contributed to experimental design and data analysis. ST contributed to experimental design and execution. MB, IBZ, JB, and NS contributed to computational work on guide design. LM contributed to cohort design and collection strategy. RB contributed to experimental design, data analysis, and editing.

