## Supplemental Information for "BADLOCK: A Rapid, Portable, Inexpensive Diagnostic for Bacterial Pathogen and Resistance Detection in Resource-Limited Settings" (from another version of this preprint: version 1, 2025-08-13)

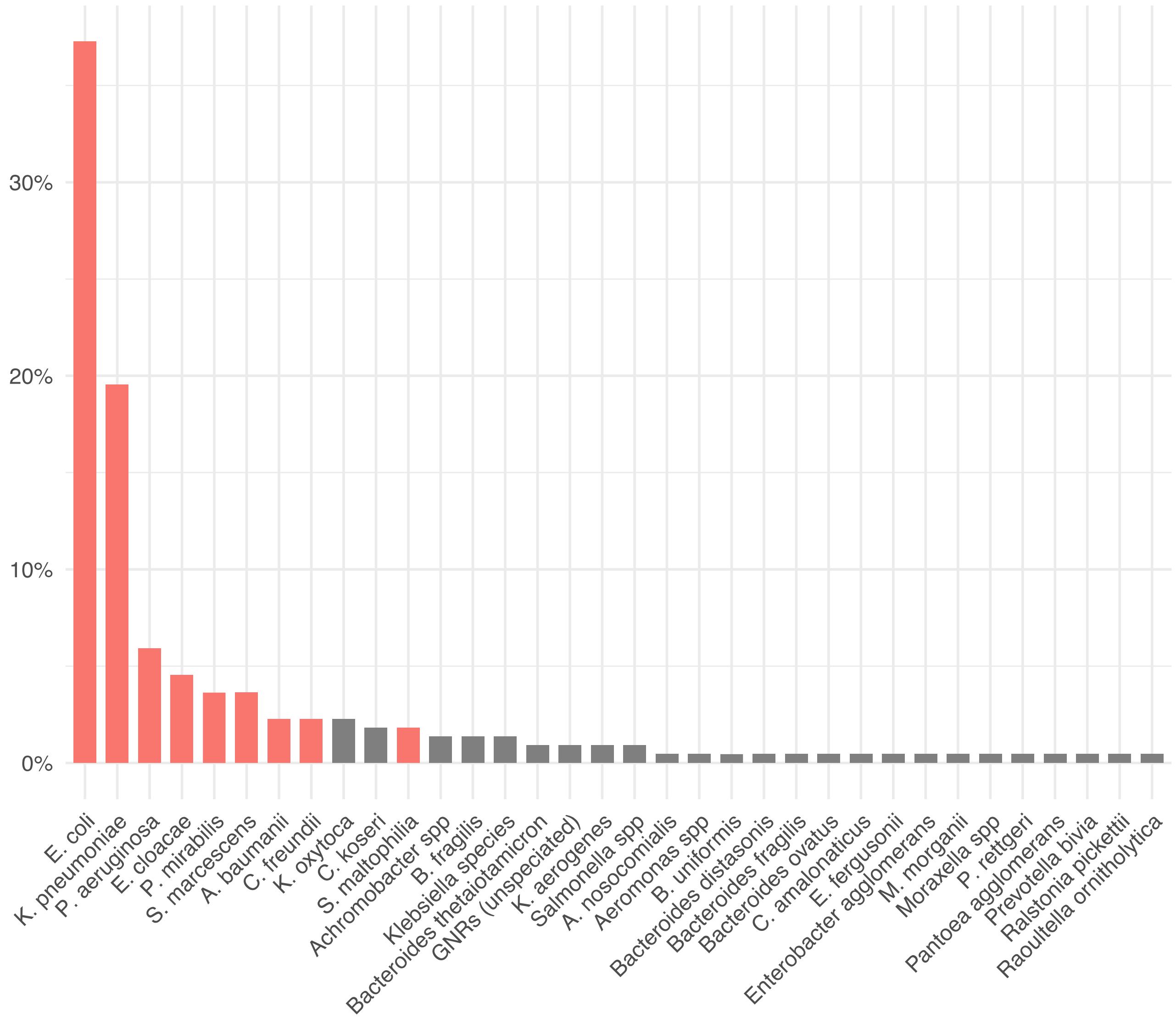


**Figure S1.** Proportional representation of gram-negative rod species from our entire clinical cohort. Species included in BADLOCK are shown in red.

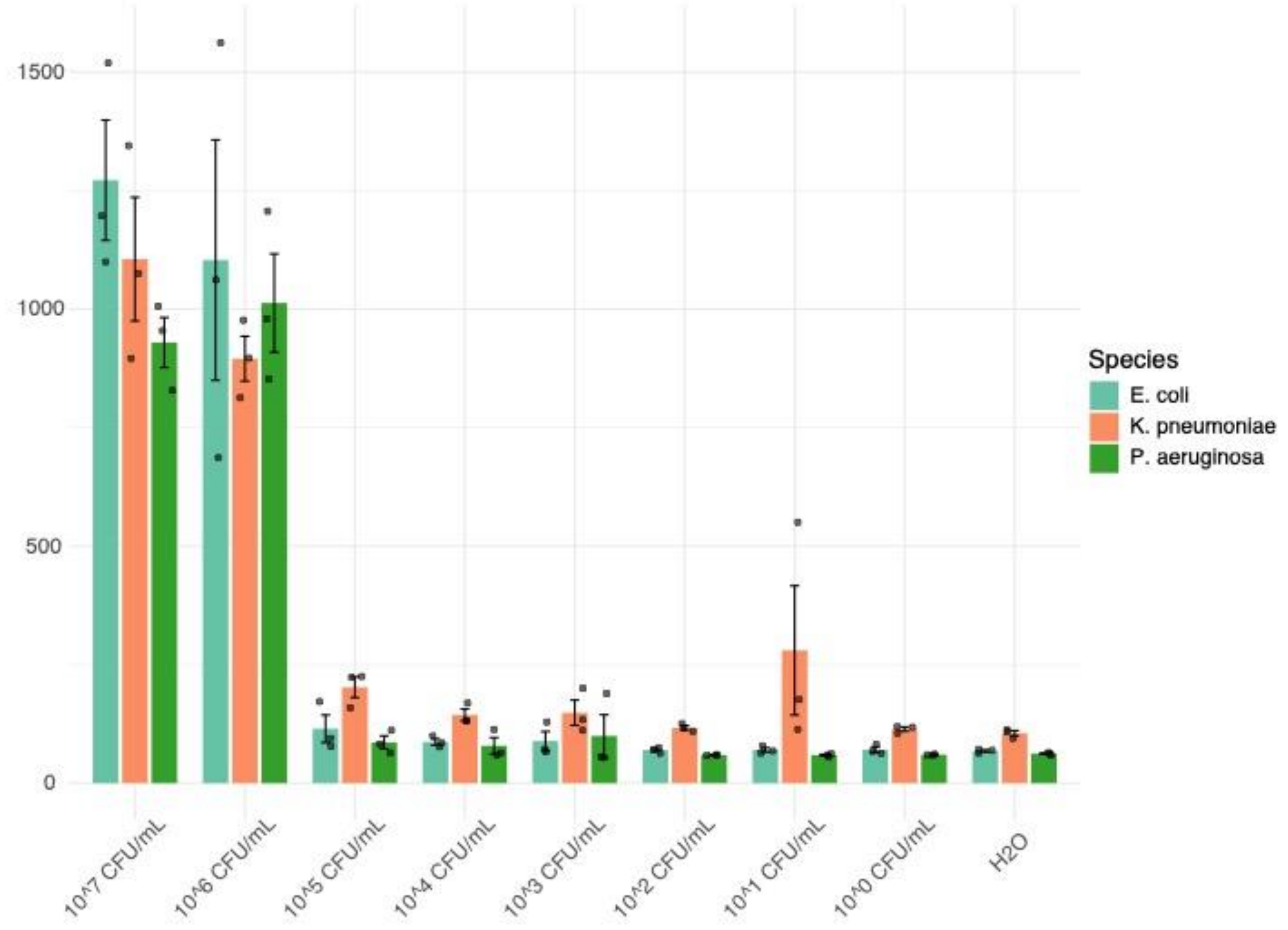


**Figure S2.** Limit of detection (LOD) analysis of the BADLOCK assay using LB-cultured isolates. RFU values are shown for three representative bacterial species across a serial dilution range measured in CFU/mL. The BADLOCK assay reproducibly detects targets at concentrations of ~$10^6$ CFU/mL (corresponding to $10^3$ CFU/uL), establishing this as the approximate lower limit of detection under these experimental conditions.

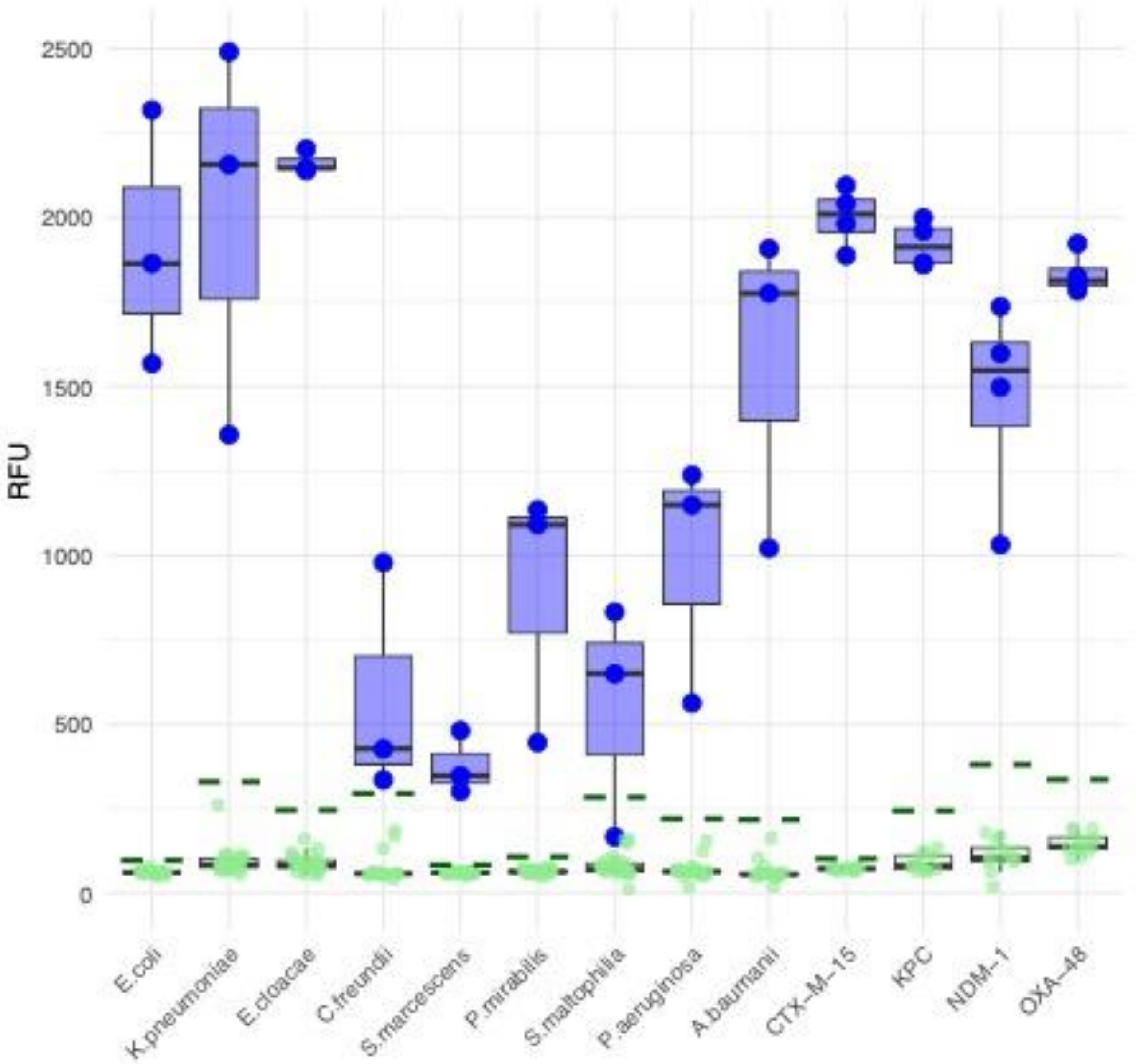


**Figure S3.** Threshold determination for species and antimicrobial resistance gene targets in the BADLOCK assay. Fluorescence values from technical replicates are shown for species targets using clinical samples, and for antimicrobial resistance (AMR) gene targets using banked clinical strains. The horizontal dashed line indicates the threshold for calling a positive result, defined as the mean plus six times the standard deviation of fluorescence values from 24 off-target samples for species reactions and 12 off-target samples for AMR gene reactions.

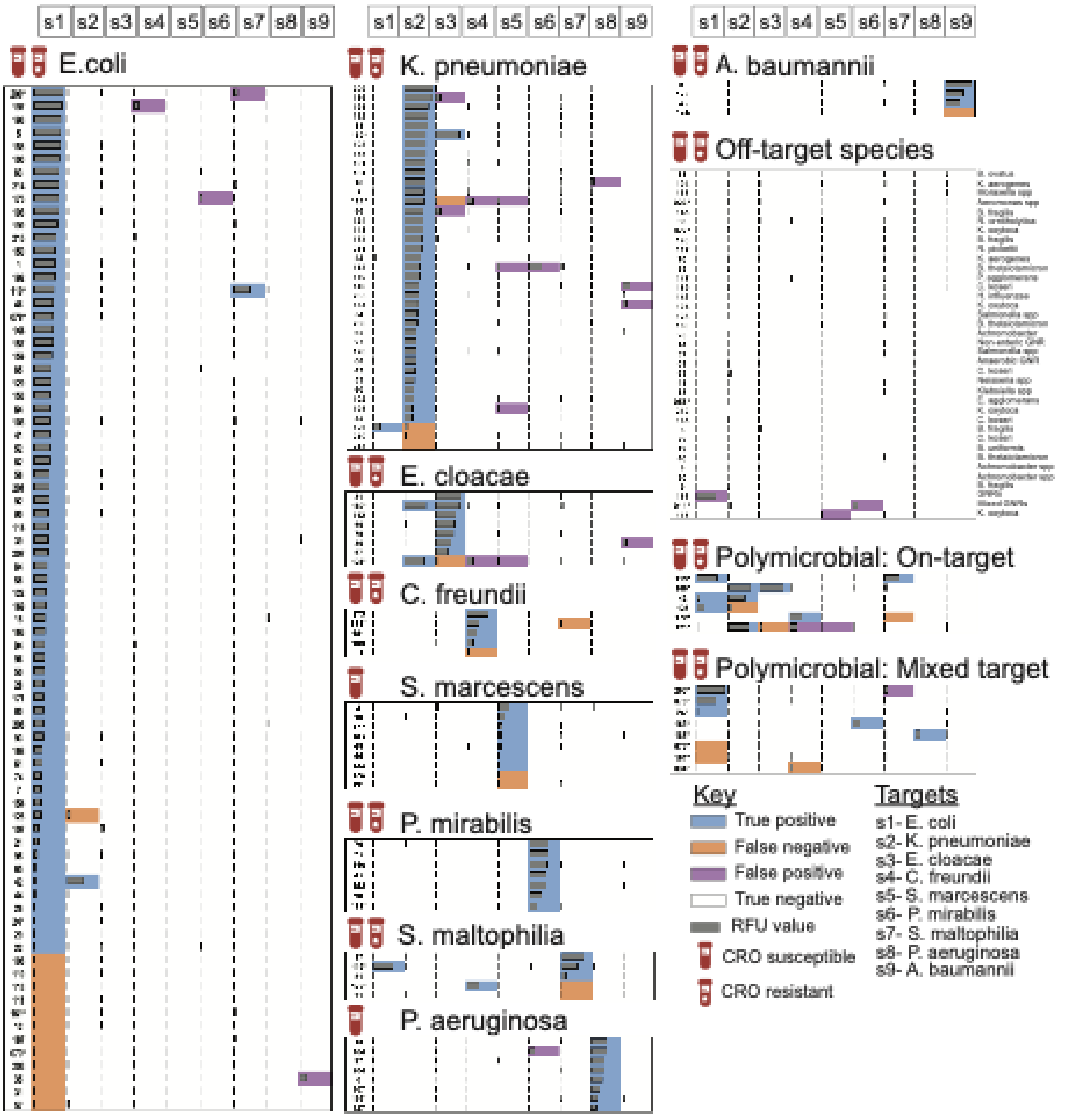


**Figure S4.** Consolidated visualization of BADLOCK species assay runs across all tested blood culture samples collected for this study. Each row represents an individual sample, and each column corresponds to a specific BADLOCK target with the assay layout shown in the boxes above and enumerated in the accompanying key. Samples are grouped by the species for which they were known positives as identified by the clinical microbiology laboratory’s standard workflow. Each cell in the grid reflects a single target reaction for a given sample, color-coded by accuracy. Overlaid grey bars represent the corresponding RFU values. Symbols adjacent to species names indicate the cohort origin of each sample and are harmonized with the legend in main-text Figure 3. Only the first run for each reaction is shown here. Polymicrobial samples appear multiple times - once for each on-panel species present, and again in the “polymicrobial” section.

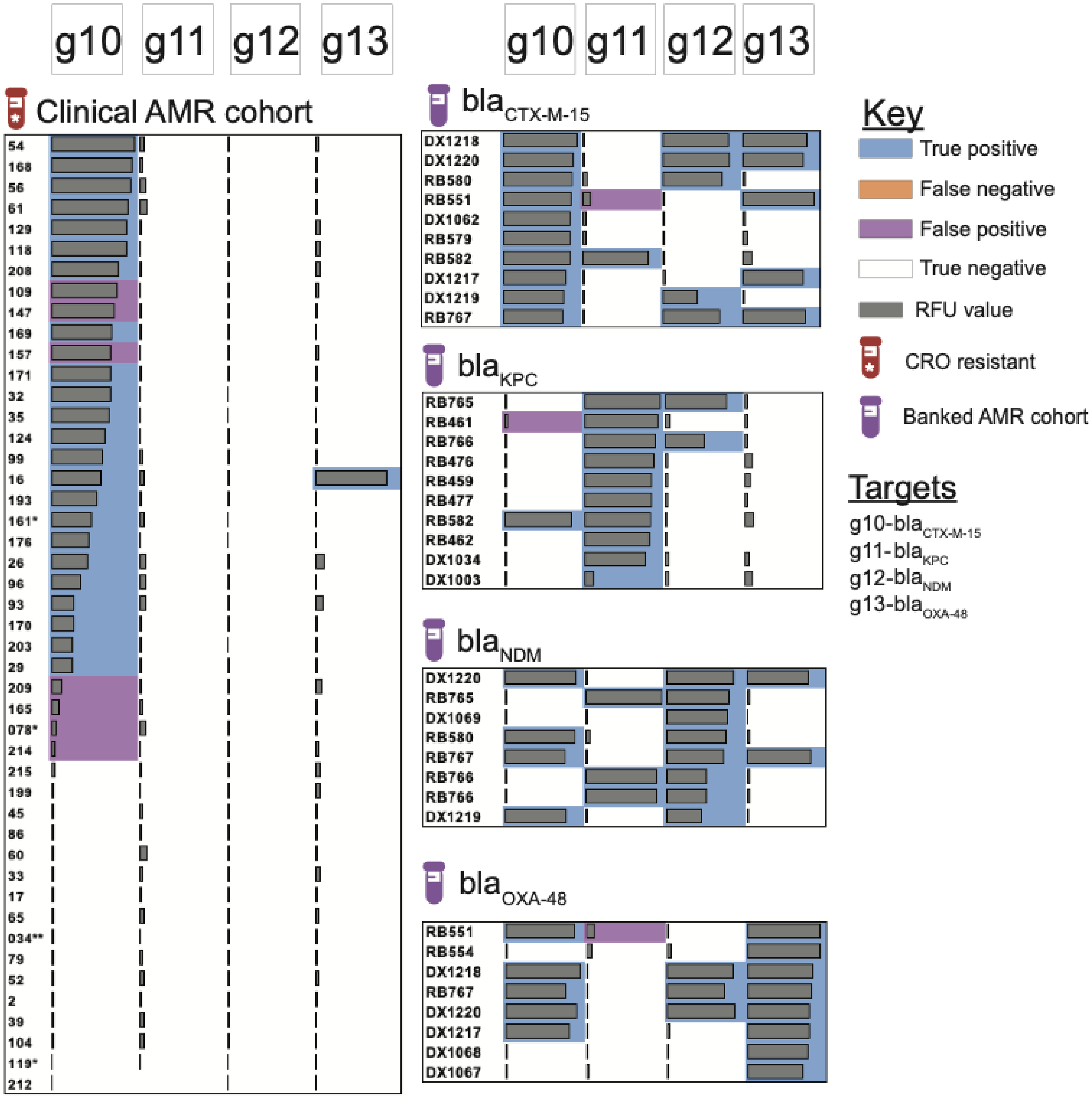


**Figure S5**. Consolidated visualization of BADLOCK AMR gene assay runs across all tested samples. Each row represents an individual sample, and each column corresponds to a specific BADLOCK AMR gene target, with the assay layout shown in the boxes above and enumerated in the accompanying key. Samples are grouped by the resistance gene known to be present by either PCR testing (clinical resistant cohort) or whole genome sequencing (banked cohort) (Table S3). Each cell in the grid reflects a single target reaction for a given sample, color-coded by accuracy, with overlaid grey bars representing the corresponding RFU values. Symbols adjacent to gene names indicate the cohort origin of each sample and are harmonized with the legend in main-text Figure 3. Samples containing multiple AMR gene targets appear in each group for which a corresponding target was known to be present.

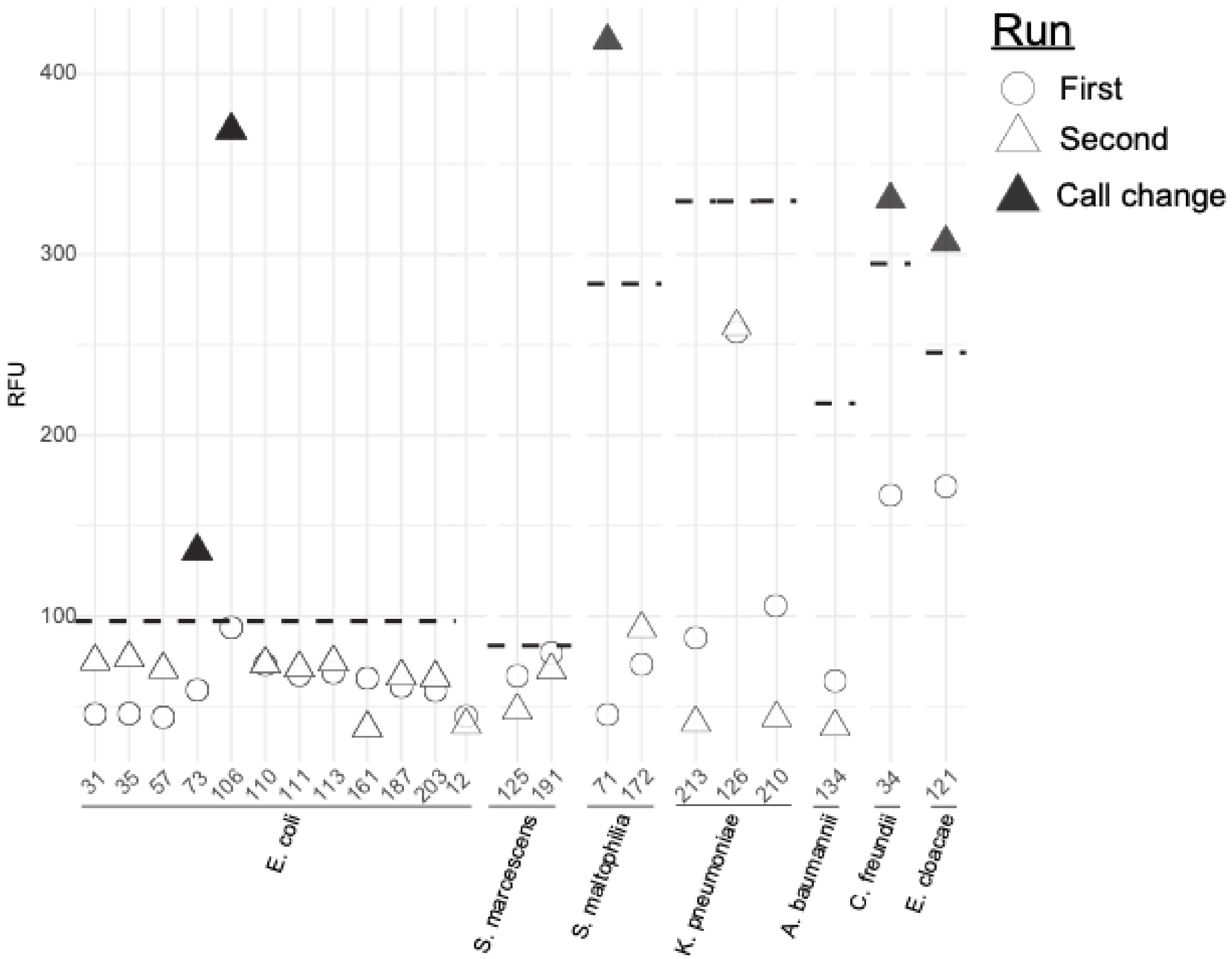


**Figure S6**. Repeat testing of falsely negative samples in the BADLOCK assay. Each point represents a clinical sample that was initially negative by BADLOCK but positive for the indicated species by the reference method. For simplicity, only the discordant targets are shown. Samples are displayed along the x-axis, grouped by species, with shape indicating whether the data correspond to the original or repeat run. The dashed horizontal line marks the species-specific RFU positivity threshold. A change in result classification occurs when a point crosses this threshold between runs, marked by a shaded triangle. 5 samples retested as concordant, while 17 remained discordant. Ten of these discordant negatives were in *E. coli* while 7 were non-*E. coli* species, and 4 of these were from polymicrobial samples (IDs 172, 126, 34, and 121). One *E. coli* sample that retested as concordant was 106, which was confirmed to have the *chuA* gene target by PCR, while the other lacked the target.

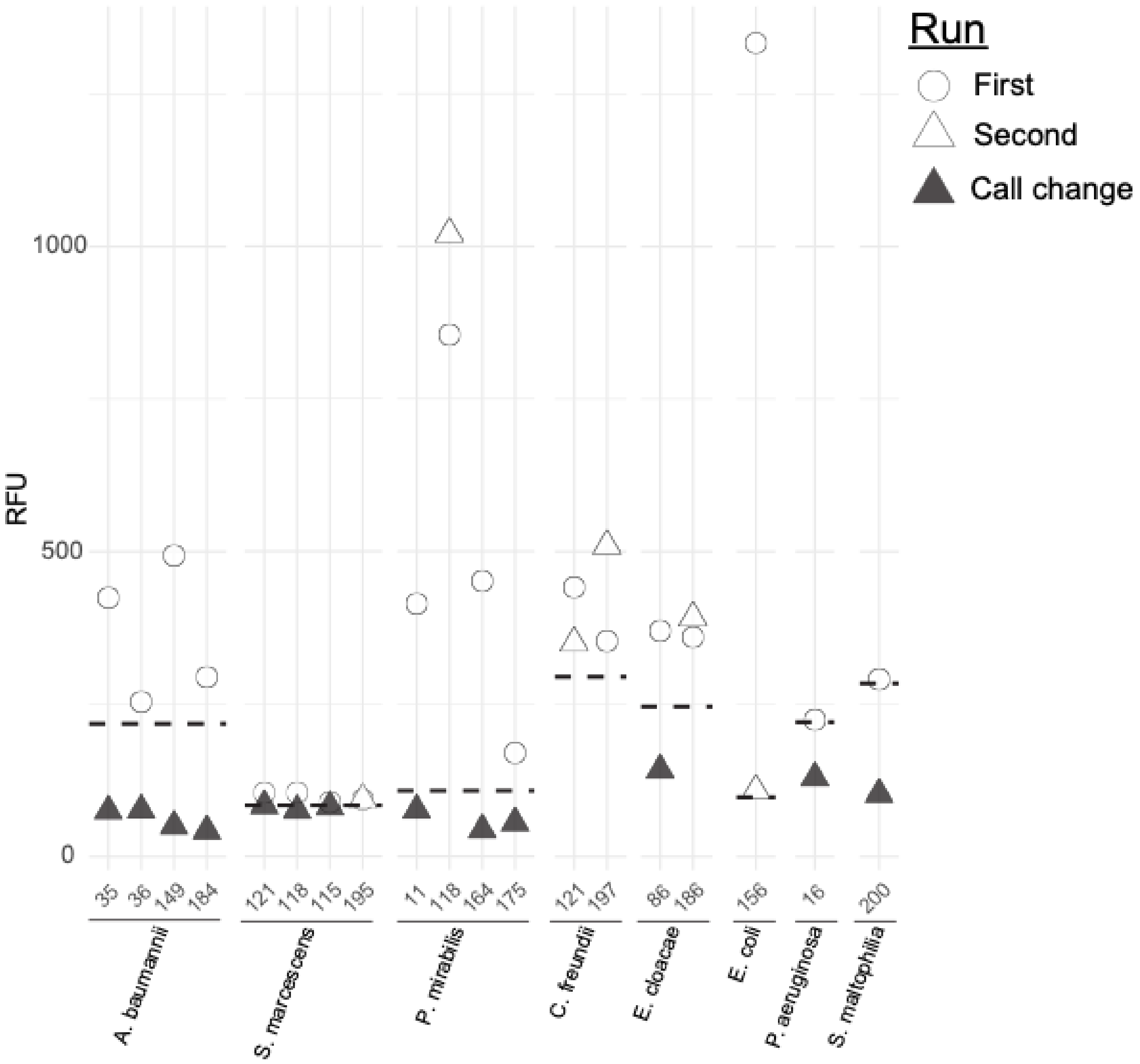


**Figure S7.** Repeat testing of discordantly positive samples in the BADLOCK assay. Each point represents a clinical sample that was initially positive by BADLOCK for the indicated species, which was not found by the reference method. For simplicity, only the discordant targets are shown. Samples are displayed on the x-axis and grouped by species, with shape distinguishing original versus repeat reactions. The dashed horizontal line indicates the species-specific RFU positivity threshold. A change in result classification occurs when a point crosses this threshold between reactions, marked by a shaded triangle. Upon retesting, 13 samples were corrected, whereas 7 remained discordant. Of these, two samples (IDs 195 and 156) yielded RFU values within 11% of the threshold in both reactions, whereas the remaining four consistently showed strong positive signals above threshold.

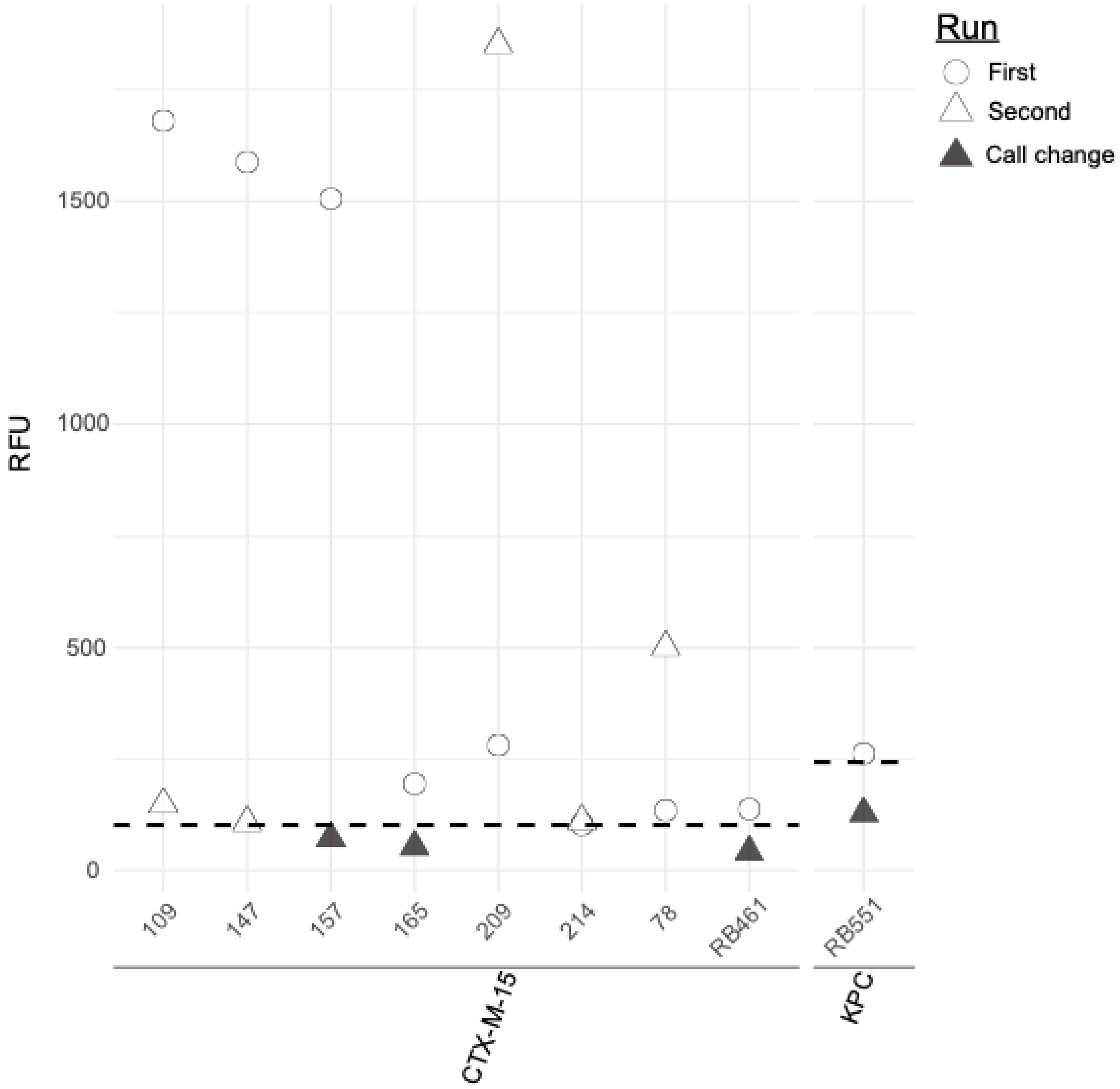


**Figure S8.** Repeat testing of discordantly positive samples in the BADLOCK AMR assay. Samples are displayed on the x-axis and grouped by AMR target, with shape distinguishing original versus repeat reactions. The dashed horizontal line indicates the target-specific RFU positivity threshold. A change in result classification occurs when a point crosses this threshold between reactions, marked by a shaded triangle.

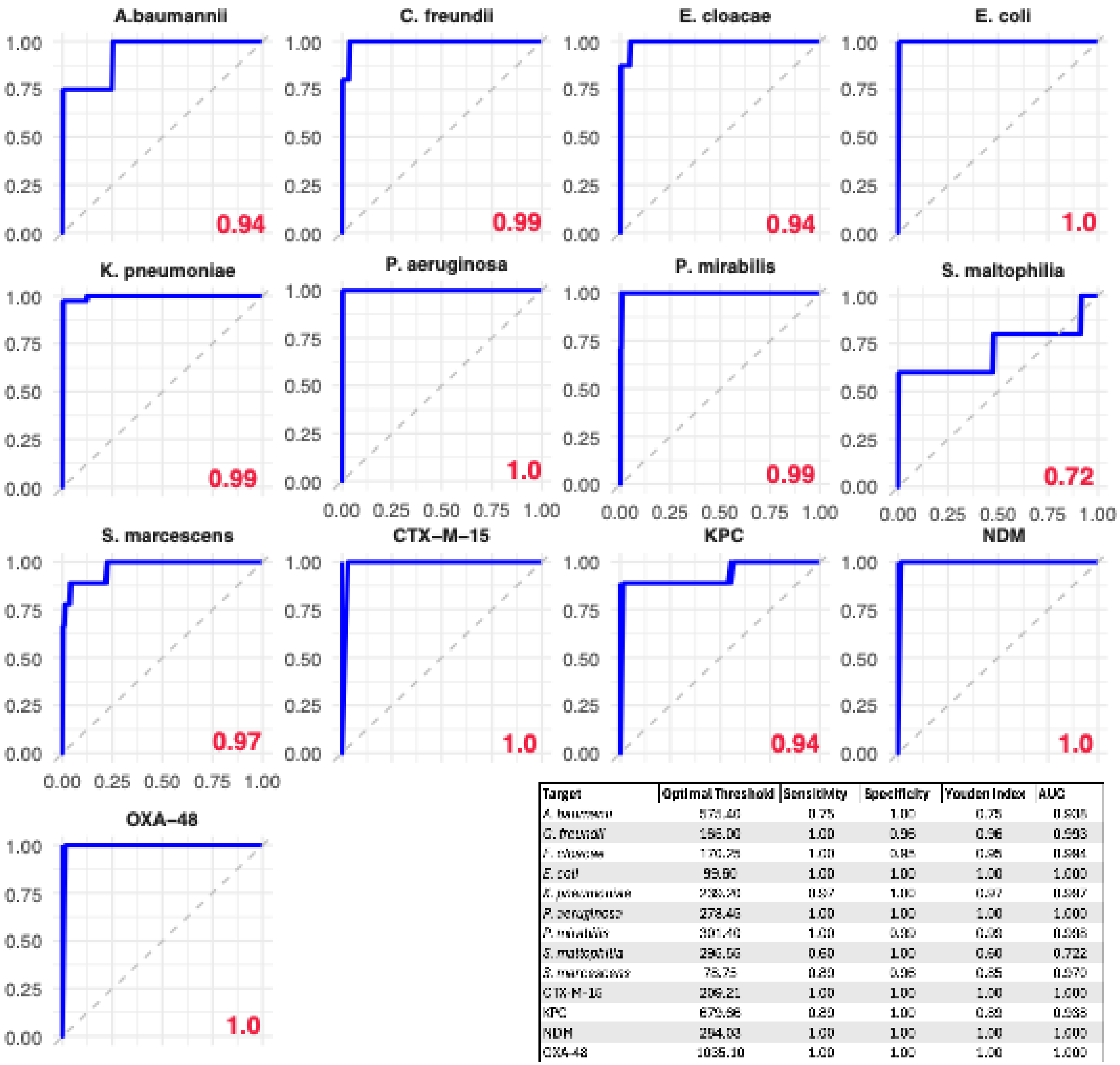


| Target | Optimal Threshold | Sensitivity | Specificity | Youden Index | AUC |
|---|---|---|---|---|---|
| *A. baumannii* | 573.40 | 0.75 | 1.00 | 0.75 | 0.938 |
| *C. freundii* | 186.00 | 1.00 | 0.96 | 0.96 | 0.993 |
| *E. cloacae* | 170.78 | 1.00 | 0.95 | 0.95 | 0.984 |
| *E. coli* | 99.90 | 1.00 | 1.00 | 1.00 | 1.000 |
| *K. pneumoniae* | 238.20 | 0.97 | 1.00 | 0.97 | 0.987 |
| *P. aeruginosa* | 278.46 | 1.00 | 1.00 | 1.00 | 1.000 |
| *P. mirabilis* | 301.40 | 1.00 | 0.98 | 0.98 | 0.988 |
| *S. maltophilia* | 296.56 | 0.60 | 1.00 | 0.60 | 0.722 |
| *S. marcescens* | 78.75 | 0.89 | 0.96 | 0.85 | 0.970 |
| CTX-M-15 | 209.21 | 1.00 | 1.00 | 1.00 | 1.000 |
| KPC | 679.96 | 0.89 | 1.00 | 0.89 | 0.938 |
| NDM | 294.03 | 1.00 | 1.00 | 1.00 | 1.000 |
| OXA-48 | 1035.10 | 1.00 | 1.00 | 1.00 | 1.000 |

**Figure S9.** Receiver operating characteristic (ROC) curves for each species-specific and AMR gene target. Curves were generated by comparing RFU values to known true-positive and true-negative classifications after excluding suspected contaminants (defined as samples with repeat RFU values >3-fold lower than the original) and *E. coli* false negatives lacking the *chuA* gene as confirmed by PCR. Each curve reflects the performance of a primer-guide target, with the area under the curve (AUC) in red text. Most targets demonstrated high discriminative ability, with AUC values ≥ 0.97 for the majority of species and AMR genes. The inset table displays the optimal threshold values along with their corresponding performance metrics across the cohort.

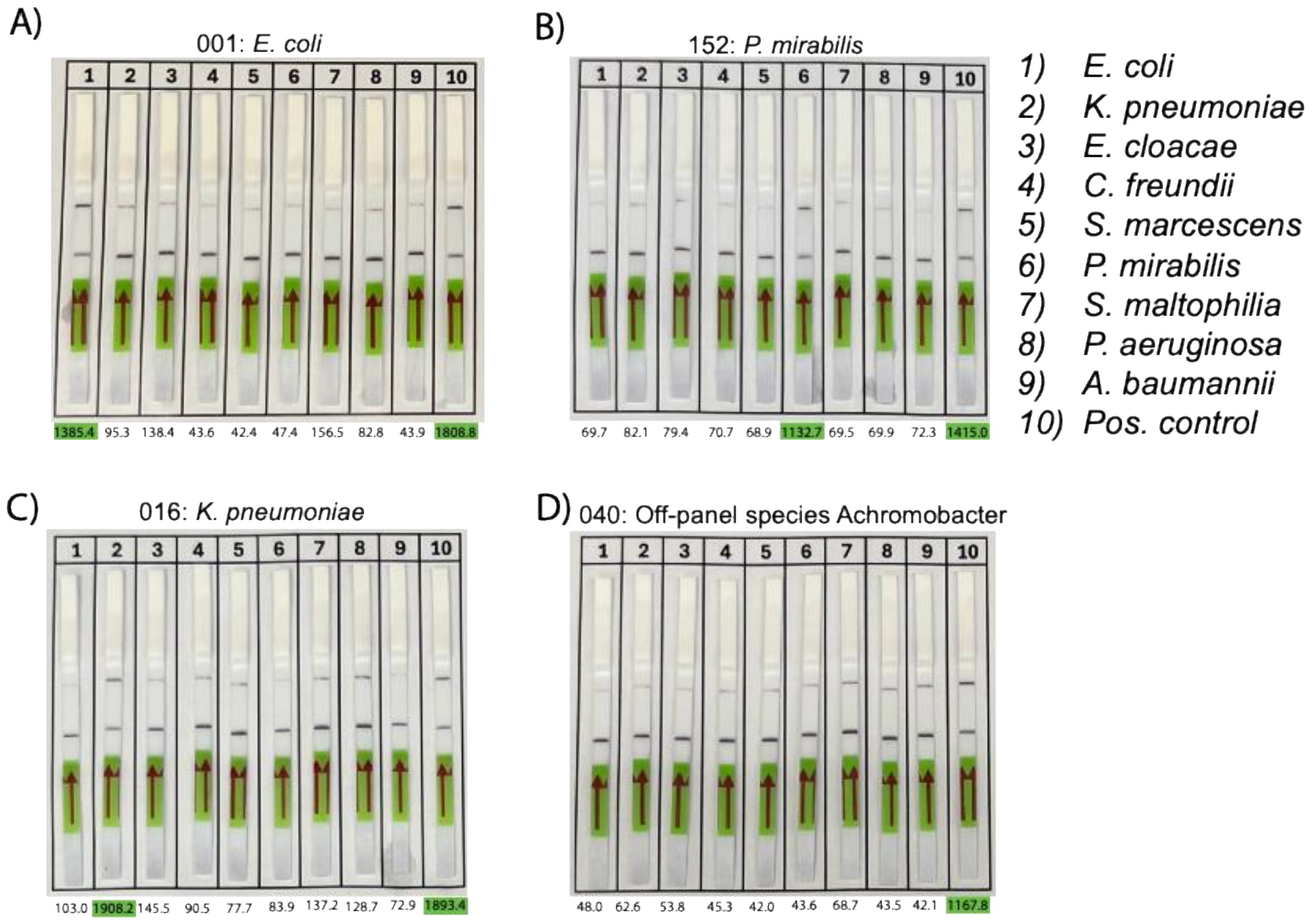


**Figure S10.** Lateral flow strips for several representative isolates, with the true species listed in the header. As in the fluorescent readout, each sample was run across all species targets in the order listed at right. RFU values from each BADLOCK reaction using the fluorescent readout are shown below, with true positive values highlighted in green.

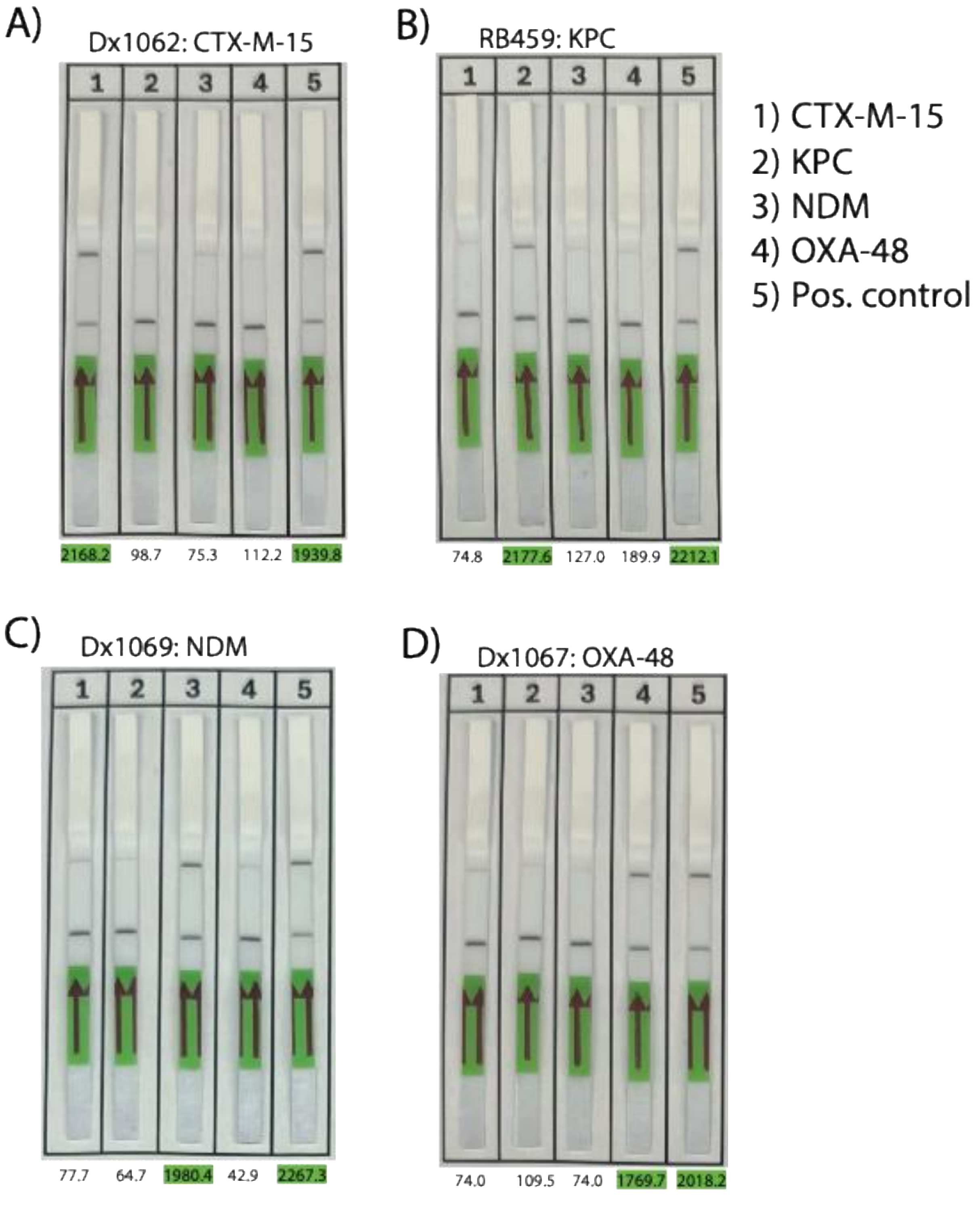


**Figure S11.** Lateral flow strips for AMR genes across different representative isolates, with the same format as above. RFU values from each BADLOCK reaction are shown below, with true positive values highlighted in green.

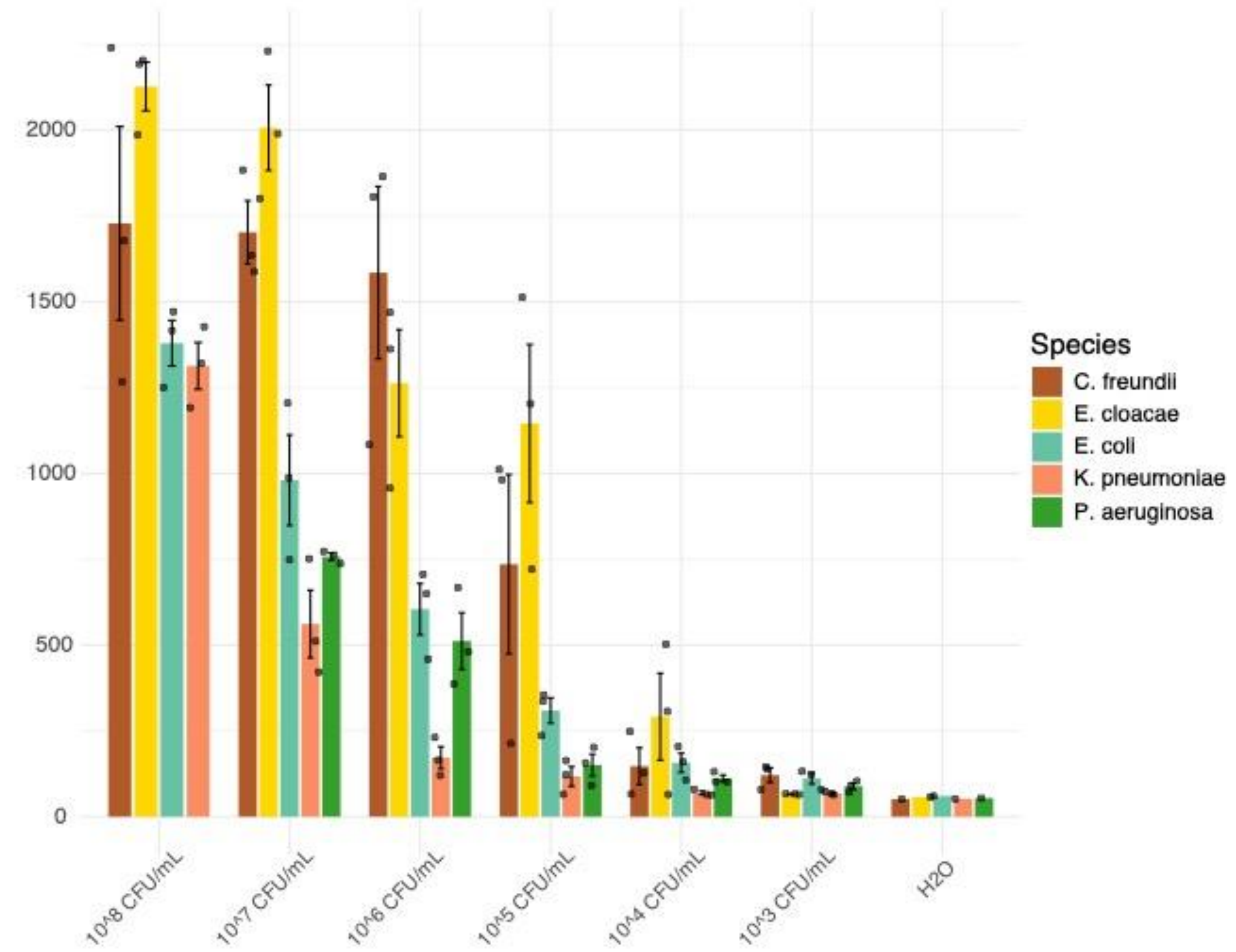


**Figure S12.** Limit of detection (LoD) for selected bacterial species included in the urine-based BADLOCK testing. Bars represent the lowest concentration (CFU/mL) at which each species was consistently detected using the BADLOCK assay. LoD values were determined using dilution series in pooled human urine, with detection defined as consistent assay positivity across replicates.

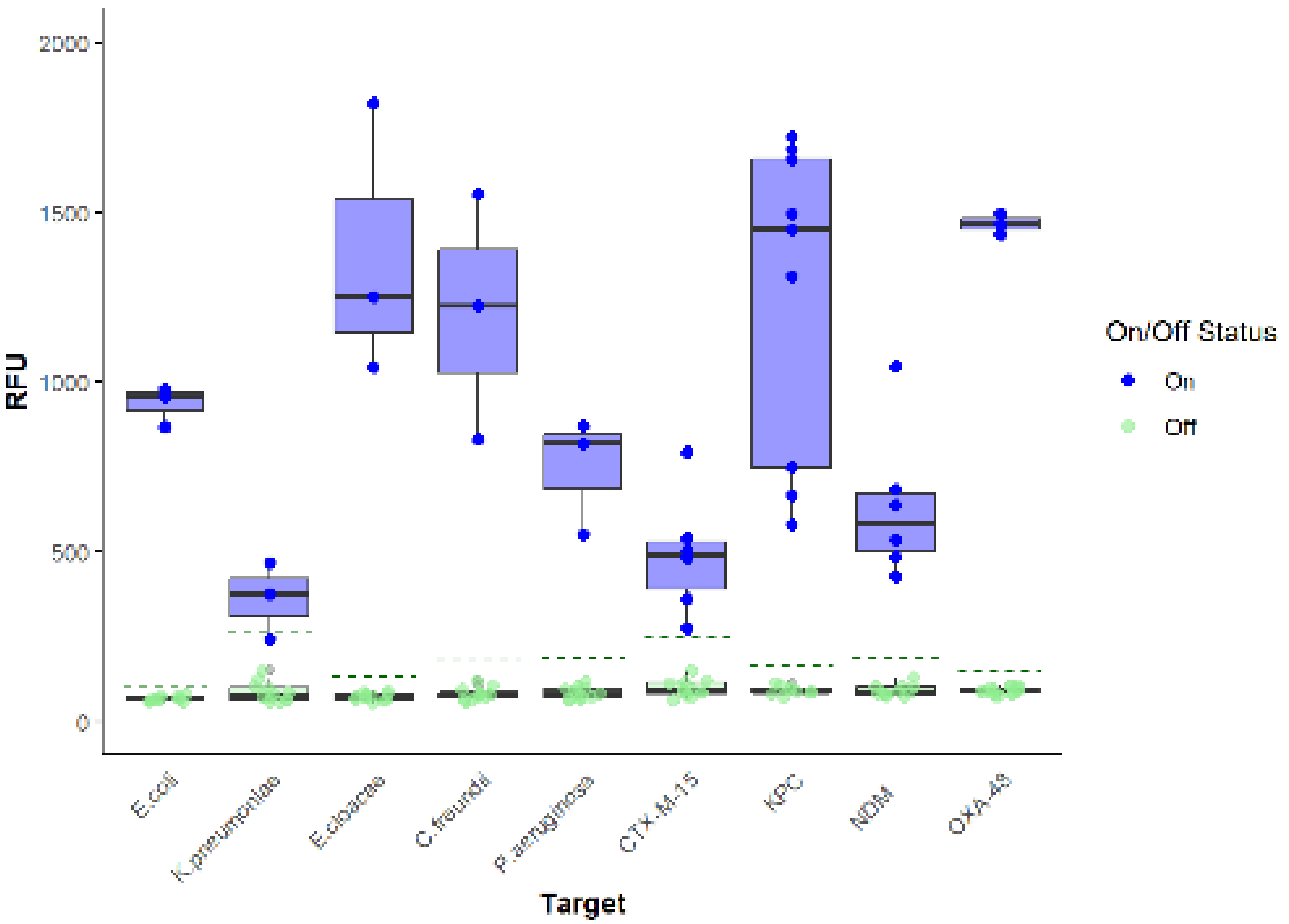


**Figure S13.** Threshold determination for species and antimicrobial resistance gene targets in the BADLOCK assay on commercial pooled human urine samples spiked with the indicated pathogen at 10^5 cfu/mL. Fluorescence values from technical replicates are shown. The horizontal dashed line indicates the threshold for calling a positive result, defined as the mean plus six times the standard deviation of fluorescence values from 12 off-target samples for species reactions and between 9 and 15 off-target samples for AMR gene reactions (variation due to some strains containing multiple target genes).

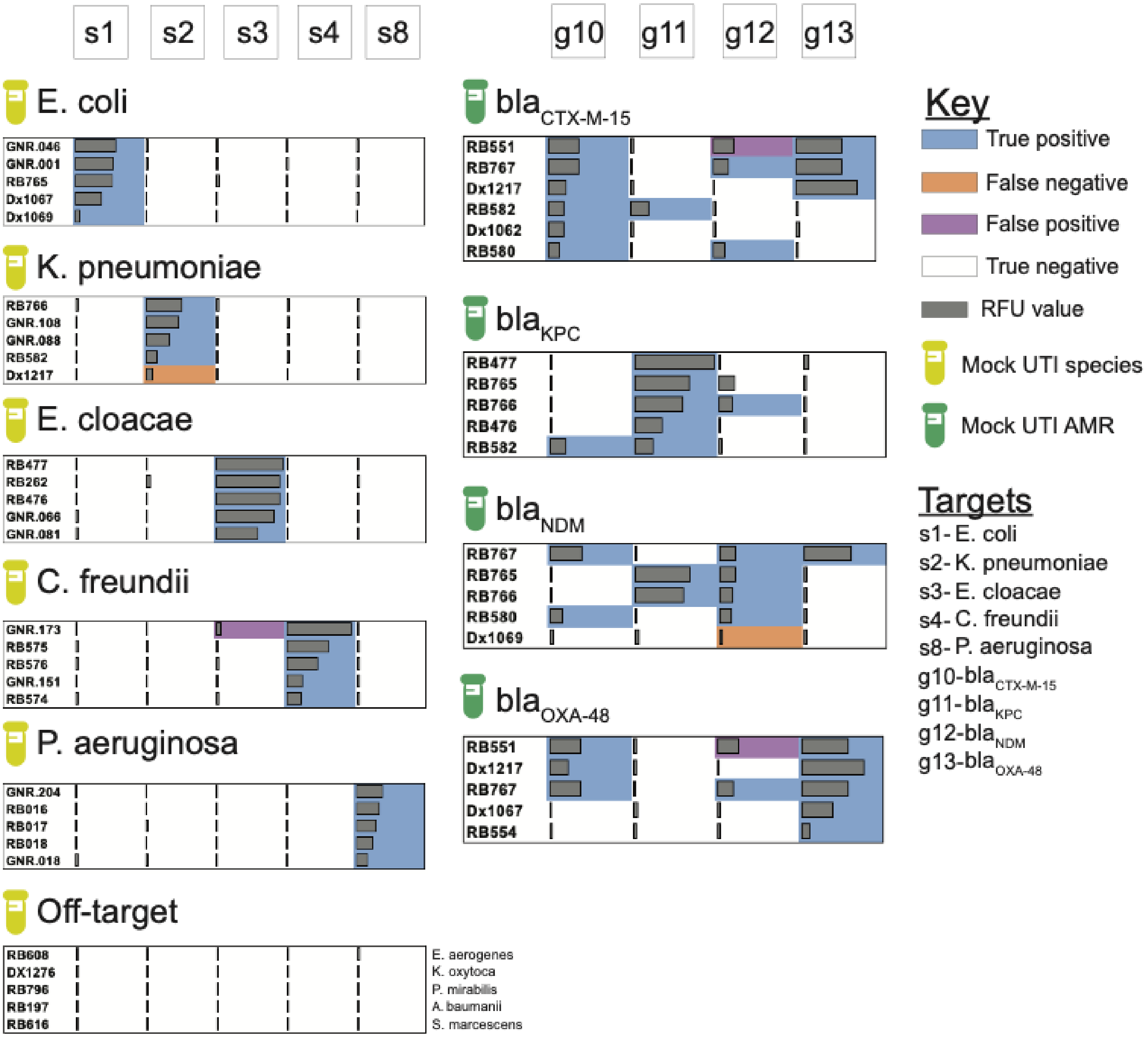


**Figure S14.** Consolidated visualization of BADLOCK assay runs for mock urinary tract infection (UTI) samples, including both species detection and AMR gene testing. Each row represents an individual sample, and each column corresponds to a BADLOCK target with the assay layout shown in the boxes above and listed in the accompanying key. Samples are grouped according to the species or AMR gene known to be present. Each cell in the grid reflects a single reaction for a given sample, color-coded by call, with overlaid grey bars representing the corresponding RFU values. Symbols adjacent to each target indicate the cohort or target class (species or resistance gene), harmonized with the legend in main-text Figure 3. Samples containing multiple targets are shown in each group for which a corresponding target was present. The off-target species are listed next to the sample row.

| Species | Target Gene | RPA Forward | RPA Reverse | Guide |
|---|---|---|---|---|
| *E. coli* | *chuA* | gaaattaatacgactcactatagggATATGGCGGTGAGTATTATCGTCAGGAACAACATC | GAGATGACCATTTGTCGGCATCAACATCTTTGTAG | CGGCGCGACGACGGGCTTCCCGCAAGCAgttttagtccccttcgtttttggggtagtctaaatcccctatagtgagtcgtattaatttc |
| *K. pneumoniae* | *topA* | gaaattaatacgactcactatagggGGTGACGCCCTGCCGCTGCAGGTAACCCATAA | ATCTTCACGCTCCAGTACGCTGTAGCTGGCTTT | GTGATGAGACCATGGCGGCGGTATCGCTgttttagtccccttcgtttttggggtagtctaaatcccctatagtgagtcgtattaatttc |
| *P. aeruginosa* | *lasB* | gaaattaatacgactcactatagggATGTTCTATCCGCTGGTGTCGCTGGA | ACCGCTGCCCTTCTTGATGTCGTAG | TTCTATATGCGCGGCAAGAACGACTTCCgttttagtccccttcgtttttggggtagtctaaatcccctatagtgagtcgtattaatttc |
| *E. cloacae* | *contig23* | gaaattaatacgactcactatagggAATGGGCGTATGATTATACCCAGGTAA | AGCAGTTTGGCAAAGTGCGCTTGTGCAG | AGTCAACTATTAGGCCAAAGCTATGATCgttttagtccccttcgtttttggggtagtctaaatcccctatagtgagtcgtattaatttc |
| *C. freundii* | *topA* | gaaattaatacgactcactatagggGTTGAAAACCGTCGCTTTTACGCTGAAAAA | TCCATTCGGCTTCTTTATTCGCCACCTGGTC | ATGGGTGAAATCGTCACCGATCGTCTGGgttttagtccccttcgtttttggggtagtctaaatcccctatagtgagtcgtattaatttc |
| *S. maltophilia* | *400kb_contig* | gaaattaatacgactcactatagggGACCATCCTGTACGCGCTGGGCTGGACGCAGC | CACCACGATTTCATCAGGCTGACGTGGAACTTC | TGTCCGACCTGCTGCCCGGCTACCTGACgttttagtccccttcgtttttggggtagtctaaatcccctatagtgagtcgtattaatttc |
| *P. mirabilis* | *topA* | gaaattaatacgactcactatagggCGGTACGCCTACTCGTCGAGCGTGAACGTG | CGTCACTTCCATACGCAAGGGTTGTTCATC | ATTGGCAATTACACGCTTCACTGCTGACgttttagtccccttcgtttttggggtagtctaaatcccctatagtgagtcgtattaatttc |
| *A. baumannii* | *topA* | gaaattaatacgactcactatagggGGCACTTACGCGAAGTGATTGGTGGTGAC | CACAGTAATGGCGAAACCATGAAGCCTAC | CAGCCAACACGTCTCGACTTAAACCGTGgttttagtccccttcgtttttggggtagtctaaatcccctatagtgagtcgtattaatttc |
| *S. marcescens* | *luxS* | gaaattaatacgactcactatagggGAAGAGCAGCGCGTTGCCGATGCCTGGAAAG | CTTTCGGCAGCGCCAGTTCGTCGTTGTGGT | TGTGGTACCTACCACATGCACTCGCTGGgttttagtccccttcgtttttggggtagtctaaatcccctatagtgagtcgtattaatttc |
| NA | KPC | gaaattaatacgactcactatagggCGTCTAGTTCTGCTGTCTTGTCTCTCATGG | CAAAGTCCTGTTCGAGTTTAGCGAATGGTT | GCTGGCTGGCTTTTCTGCCACCGCGCTGgttttagtccccttcgtttttggggtagtctaaatcccctatagtgagtcgtattaatttc |
| NA | NDM | gaaattaatacgactcactatagggAATGTCTGGCAGCACACTTCCTATCTCGAC | TGATCTCCTGCTTGATCCAGTTGAGGATCT | CAACGGTTTGATCGTCAGGGATGGCGGCgttttagtccccttcgtttttggggtagtctaaatcccctatagtgagtcgtattaatttc |
| NA | CTX-M-15 | gaaattaatacgactcactatagggATAAAACCGGCAGCGGTGGCTATGG | GCTAATACATCGCGACGGCTTTCTGCCTTA | ATCGTGCGCCGCTGATTCTGGTCACTTAgttttagtccccttcgtttttggggtagtctaaatcccctatagtgagtcgtattaatttc |
| NA | OXA-48 | gaaattaatacgactcactatagggTTAAAATTCCCAATAGCTTGATCGCCCTCG | ATAAACAGGCACAACTGAATATTTCATCGC | CCAAGTCTTTAAGTGGGATGGACAGACGgttttagtccccttcgtttttggggtagtctaaatcccctatagtgagtcgtattaatttc |
| Synthetic target | NA | gaaattaatacgactcactatagggg | TAGTTATTGCTCAGCGGTGG | GGCCGTCCGTTAATTTCCCTTGCATACAgttttagtccccttcgtttttggggtagtctaaatcccctatagtgagtcgtattaatttc |

**Table S1.** Primer and guide sequences for each target gene included in our panel. Lowercase letters indicate conserved sequences (T7 primer sequence for the forward RPA primer, and non-targeting region of the guide sequence, respectively), whereas uppercase are the target-specific component.

| Species | True (+) | False (+) | True (-) | False (-) | Sensitivity | Specificity |
|---|---|---|---|---|---|---|
| *E. coli* | 66 | 1 | 115 | 12 | 84.6 | 99.1 |
| *K. pneumoniae* | 36 | 0 | 155 | 3 | 92.3 | 100 |
| *E. cloacae* | 7 | 2 | 184 | 1 | 87.5 | 98.9 |
| *C. freundii* | 4 | 2 | 187 | 1 | 80 | 98.9 |
| *S. marcescens* | 7 | 4 | 181 | 2 | 77.8 | 97.8 |
| *P. mirabilis* | 7 | 4 | 183 | 0 | 100 | 97.9 |
| *S. maltophilia* | 3 | 1 | 188 | 2 | 60 | 99.5 |
| *P. aeruginosa* | 8 | 1 | 185 | 0 | 100 | 99.5 |
| *A. baumannii* | 3 | 4 | 186 | 1 | 75 | 97.9 |

**Table S2.** Performance characteristics for each species target.

| Sample ID | Published ID | Source of Strain | Cohorts | Species | AMR genes |
|---|---|---|---|---|---|
| DX1003 | MGH145 | PMID: 28096418 | AMR | *C. freundii* | KPC-2 |
| DX1034 | DH10Beta + pBAD_KPC3 | Bhattacharyya Lab Strain | AMR | *E. coli* | KPC-3 |
| DX1062 | RB001+CTX-M-15 | Bhattacharyya Lab Strain | AMR, Urine species | *E. coli* | CTX-M-15 |
| DX1067 | RB001+OXA-48 | Bhattacharyya Lab Strain | AMR, Urine species, Urine AMR | *E. coli* | OXA-48 |
| DX1068 | RB059+OXA-48 | Bhattacharyya Lab Strain | AMR | *E. coli* | OXA-48 |
| DX1069 | RB001+NDM-1 | Bhattacharyya Lab Strain | AMR, Urine species, Urine AMR | *E. coli* | NDM-1 |
| DX1217 | AR0066 (CDC) | CDC AR Isolate Bank | AMR, Urine species, Urine AMR | *K. pneumoniae* | CTX-M-15, OXA-232 |
| DX1218 | AR0068 (CDC) | CDC AR Isolate Bank | AMR | *K. pneumoniae* | CTX-M-15, NDM-1, OXA-48 |
| DX1219 | AR0143 (CDC) | CDC AR Isolate Bank | AMR | *K. pneumoniae* | CTX-M-15, NDM-1 |
| DX1220 | AR0153 (CDC) | CDC AR Isolate Bank | AMR | *K. pneumoniae* | CTX-M-15, NDM-1, OXA-48 |
| DX1276 | MGH175 | PMID: 28096418 | Urine species | *K. oxytoca* | na |
| RB016 | 2732233 | Bhattacharyya Lab Strain | Urine species | *P.aeruginosa* | na |
| RB017 | B895023 | Bhattacharyya Lab Strain | Urine species | *P.aeruginosa* | na |
| RB018 | 2762085 | Bhattacharyya Lab Strain | Urine species | *P.aeruginosa* | na |
| RB197 | Acinetobacter #1 | Bhattacharyya Lab Strain | Urine species | *A. baumanii* | na |
| RB262 | SBJ-32413; 1349 | Bhattacharyya Lab Strain | Urine species | *E. cloacae* | na |
| RB459 | BIDMC68 | PMID: 28096418 | AMR | *K. pneumoniae* | KPC-2 |
| RB461 | MGH67 | PMID: 28096418 | AMR | *K. pneumoniae* | KPC-2 |
| RB462 | MGH71 | PMID: 28096418 | AMR | *K. pneumoniae* | KPC-2 |
| RB476 | BWH39 | PMID: 28096418 | AMR, Urine species, Urine AMR | *E. cloacae* | KPC-2 |
| RB477 | MGH5 | PMID: 28096418 | AMR, Urine species, Urine AMR | *E. cloacae* | KPC-2 |
| RB551 | BWH2 | PMID: 28096418 | AMR, Urine AMR | *K. pneumoniae* | CTX-M-15, OXA-48 |
| RB554 | BAA2524 | PMID: 28096418 | AMR, Urine AMR | *K. pneumoniae* | OXA-48 |
| RB574 | MGH281c | PMID: 28096418 | Urine species | *C. freundii* | na |
| RB575 | MGH281 | PMID: 28096418 | Urine species | *C. freundii* | na |
| RB576 | MGH283 | PMID: 28096418 | Urine species | *C. freundii* | na |
| RB579 | IDR1600031102-01-00 | Wadsworth Lab | AMR | *K. pneumoniae* | CTX-M-15 |
| RB580 | IDR1600014966-01-00 | Wadsworth Lab | AMR, Urine AMR | *Citrobacter spp* | CTX-M-15, NDM-1 |
| RB582 | IDR1600037319-01-00 | Wadsworth Lab | AMR, Urine species, Urine AMR | *K. pneumoniae* | CTX-M-15, KPC-2 |
| RB608 | BAC0800002538 | Wadsworth Lab | Urine species | *E. aerogenes* | na |
| RB616 | IDR1300010999 | Wadsworth Lab | Urine species | *S. marcescens* | na |
| RB765 | F3461517 | PMID: 28096418 | AMR, Urine species, Urine AMR | *E. coli* | KPC-2, NDM-1 |
| RB766 | F3321027 | PMID: 28096418 | AMR, Urine species, Urine AMR | *K. pneumoniae* | KPC-2, NDM-1 |
| RB767 | X1621267 | PMID: 28096418 | AMR, Urine AMR | *E. coli* | CTX-M-15, NDM-1, OXA-48 |
| RB796 | MGH013 | PMID: 28096418 | Urine species | *P. mirabilis* | na |

**Table S3.** Banked clinical isolates with known genomic content. The table includes the testing cohort(s) in which each sample was used, the species name, and the relevant antimicrobial resistance gene(s) of interest.
